# Evaluation of out-of-hospital management of asthma attacks prior to consultation at the paediatric emergency department in Martinique

**DOI:** 10.64898/2026.11.13.26368029

**Authors:** Alexis Julia, Julien Denis

## Abstract

**Background:** Asthma is a chronic, multifactorial disease that progresses in episodes and requires effective coordination of care between healthcare professionals and patients.

**Objective:** To describe and evaluate pre-hospital therapeutic management of asthma attacks.

**Methods:** Cross-sectional, prospective, single-centre, observational and descriptive study of patients consulting for asthma attacks at the Fort de France paediatric emergency department between July and November 2024. All patients included were children between the ages of 1 and 18 who had already been diagnosed with asthma by a doctor and who presented to the emergency department for an asthma attack. Several other parameters were assessed, such as the parents’ perception of the emergency situation, the child’s asthma control in the month prior to the attack, the child’s usual medical follow-up, and their usual treatment.

**Results:** 175 patients were included in the study. Eight patients were excluded from the study because their questionnaire had a response rate of less than 50%.

Of the 175 patients included, only 6.7% received perfect pre-hospital care, 29.3% received good quality care, 36% received moderate quality care, and 10.3% received poor quality care before arriving at the emergency department. (17.7% did not complete all the items necessary to calculate the score).

11.6% of children did not receive treatment before consulting the emergency department. Among the 88.9% who took short-acting β2 agonists, only 52% received at least 3 series and 66.2% were given doses appropriate for their weight. 21.6% received oral corticosteroids before arriving at the emergency department.

**Conclusions:** The management of asthma attacks outside of hospital settings is not optimal for a large proportion of patients in the study. Several areas for improvement have been identified and require joint action by healthcare professionals.

## INTRODUCTION

### 1 - Epidemiology

Asthma is a chronic respiratory disease with a high prevalence in the paediatric population. According to the World Health Organization (WHO), there are currently an estimated 262 million patients with asthma, and the estimated number of asthma-related deaths worldwide is 455,000 per year. (1)

In France, asthma affects 4 million people and it is estimated that 10 to 12% of the population will have asthma during their lifetime. It currently causes more than 60,000 hospitalisations and 900 deaths per year. (2) Among these deaths, around twenty are children.

Current epidemiological data show a gradual increase in the prevalence of asthma, but a decrease in mortality in France. The prevalence of asthma in children in the French West Indies is estimated at 15 to 20%, compared with 10 to 12% for children in mainland France, all regions combined. (3) This difference has not yet been clearly explained, although several hypotheses have been put forward, including higher humidity levels conducive to mould growth and the sandstorm phenomenon, which is well known in the Caribbean. A medical thesis completed in Guadeloupe in 2011 shows a correlation between sandstorm episodes and an increase in consultations for asthma exacerbations. (4)

### 2 - Clinical aspects

Asthma is a chronic inflammatory respiratory disease that manifests itself in attacks of varying intensity, which may resolve spontaneously or lead to severe respiratory distress and even death.

Clinically, it is defined by a combination of several clinical signs: wheezing, dyspnoea, coughing, chest tightness and limited expiratory airflow. The clinical presentation varies, particularly in terms of symptom intensity, depending on the attack and the individual.

The diagnosis is based primarily on the combination of symptoms presented by the patient and their progression over time. It will be confirmed by several paraclinical tests carried out outside of attacks (respiratory function testing, peak flow, stress test), in order to make a definitive diagnosis and refine the therapeutic management.

The clinical expression of asthma is highly variable, and asthma is currently classified into different clinical-biological phenotypes: (5)

● eosinophilic asthma

● neutrophilic asthma

● asthma with predominant cough

● late-onset asthma (first symptoms in adulthood)

● asthma with persistent ventilatory limitation

● asthma with obesity

### 3 - Risk factors

The factors contributing to asthma attacks are numerous, multifactorial and sometimes poorly understood. They make it difficult for health and public health authorities to control and prevent asthma attacks in the general population.

These include allergens, irritants (such as tobacco smoke) (6), air pollutants (such as fine particles), intense physical activity, climates (humid and hot), and viral respiratory infections (RSV/adenovirus, etc.). In certain situations, several interrelated risk factors can lead to an attack, hence the importance of isolating the causative agents in order to adapt the environment and lifestyle habits of patients.

Numerous genes that may contribute to asthma have been isolated, particularly in certain regions of chromosome 17 (7), however, this knowledge is limited to the field of research and its incorporation into clinical practice is currently limited. There is no organised screening for asthma to date.

### 4 - Evolution

Asthma most often manifests itself in childhood. It can persist into adulthood, manifest itself subclinically, or sometimes fade over time. (8)

The impact on the quality of life of patients and their loved ones (particularly parents of children with asthma) is significant.

The frequency and intensity of exacerbations can lead to repeated visits to healthcare facilities. (9) Exacerbations manifest as respiratory distress, which can sometimes be severe, with symptoms that are often very noisy and anxiety-inducing for the patient and those accompanying them. This can lead to repeated visits to emergency departments and a deterioration in quality of life in the short to medium term.

The repercussions can be long-term, as there is a close link between the severity of asthma in childhood and the risk of developing chronic obstructive pulmonary disease in adulthood. (10)

To date, asthma-related morbidity remains high, and despite advances in treatment and support, asthma is a public health priority in France and around the world.

### 5 - Crisis treatments

Short-acting β2 agonists (SABAs) are the cornerstone of emergency management. Their use is standardised and the dose administered will depend on the clinical severity of the attack and the patient’s weight. (11)

The recommendations of the SP2A (Pediatric Pulmonology and Allergology Society) define the management of moderate to severe asthma attacks as appropriate if:

❖ Administration of short-acting β2 agonists

❖ Adjusted to the patient’s weight (4 to 12 puffs per series)

❖ At least 3 series administered

❖ 20-minute interval between SABA series

❖ Oral corticosteroids administered from the 3rd SABA series onwards

❖ Call the emergency number if symptoms persist and signs of severity are present

The administration of SABA requires the use of an inhalation chamber with a mask (for children aged 0 to 3 years) or a mouthpiece (for children aged 4 to 12 years).

Scores are used by medical teams to assess the severity of an attack. Several scores have been validated to date, including the Asthma Severity Score, PRAM and Pulmonary Index Score, and their use depends on the centre. The PRAM score is currently used in the Martinique pediatrics emergency department. (appendix 1) It includes several clinical warning signs that parents should look out for.

A moderate asthma attack (PRAM 4) or one with a risk of complications according to the recommendations, the following treatment should be given:

❖ Inhale four to twelve puffs of SABA adjusted to the patient’s weight, every twenty minutes three times, then every one to four hours if symptoms improve.

❖ If symptoms persist after the third course, oral corticosteroids should be prescribed at a dosage appropriate for the patient’s weight, not exceeding 60 mg/day of prednisolone.

❖ If symptoms persist after the third series, or if there are signs of respiratory distress or clinical severity criteria, call the emergency services.

For mild attacks (PRAM 1 to 3), with isolated coughing or mild symptoms, recommendations are not fixed and are often left to the discretion of the practitioner. One possible treatment option is as follows:

❖ Inhale four puffs of SABA, repeating every twenty minutes if necessary.

❖ If symptoms worsen, follow the protocol for moderate or severe asthma attacks.

According to the GINA 2024 guidelines, an alternative treatment is preferred for patients aged 12 years and older:

❖ The combination of inhaled corticosteroids and formoterol would be preferred for initiating maintenance treatment and could be used during attacks in patients receiving this maintenance treatment.

❖ The maximum number of inhalations recommended before seeking emergency medical attention is height in one day.

The goal is to limit the number of treatments patients need to take and to promote patient adherence to treatment.

### 6 - Chronic treatments

The first-line chronic treatments are inhaled corticosteroids (ICS). They are administered daily and their dosage and use of the inhalation device must be reassessed regularly. Their use reduces asthma-related mortality, the number of hospitalizations, the frequency and severity of asthma attacks. They also improve symptom control and quality of life, and preserve respiratory function. (12)

Additional treatments may be prescribed for asthma that is not controlled by ICS alone. ICS may be combined with long-acting bronchodilators (LABA), anticholinergics, or leukotriene antagonists (LA).

For asthma that is resistant to high doses of ICS (step 5), biotherapies may be initiated in specialist centers in accordance with recent recommendations from the French Pediatric Pulmonology and Allergology Society.(11)

According to the GINA 2024 guidelines, an alternative treatment is preferred for patients aged 12 years and older, although the ICS +/- SABA option remains among the treatment options.

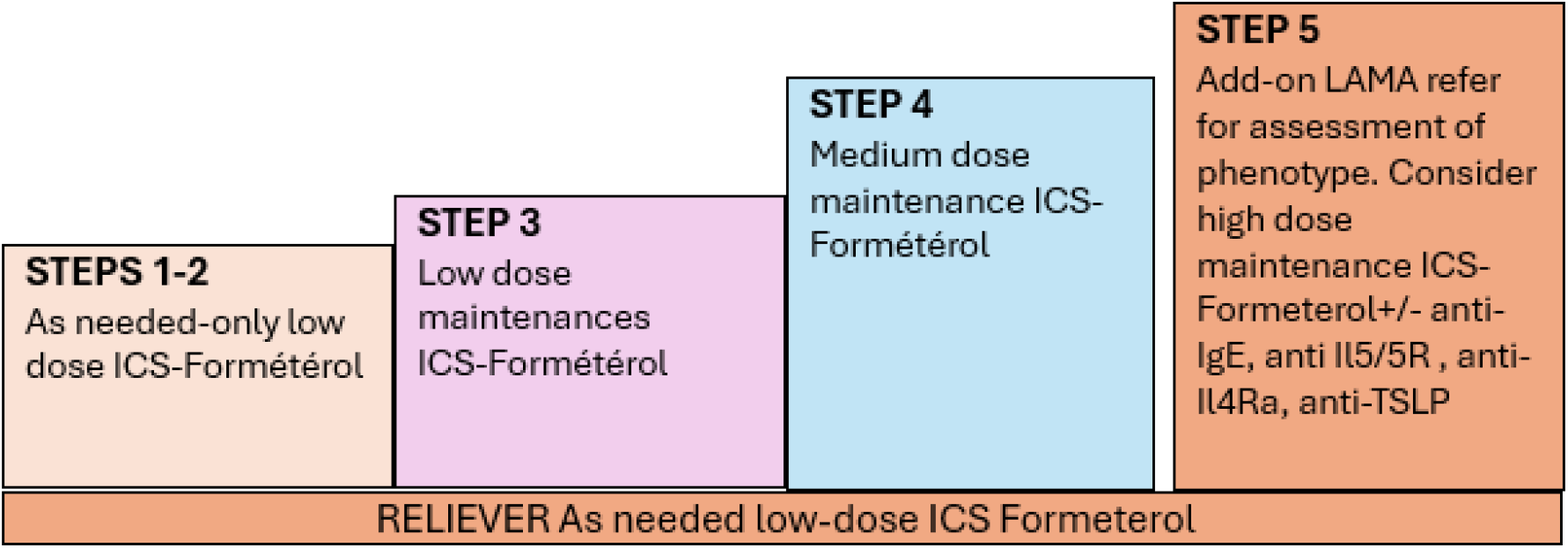

### 7 - Healthcare services in Martinique

The management of asthma requires effective coordination between various medical and paramedical professionals.

The general practitioner plays a central role in the overall management of children with asthma, but various other healthcare professionals are needed to ensure proper follow-up. Specialists (pulmonologists, pediatric pulmonologists, allergists) have a role to play, particularly in cases of moderate to severe asthma requiring more intensive treatment. Pediatric emergency services are necessary when attacks are poorly controlled despite outpatient treatment. Asthma education in hospital through the asthma school, plays a vital role in prevention and teaching children and their parents about asthma.(13)

Several obstacles to the proper monitoring of asthma patients currently exist in Martinique and France. Healthcare provision is an integral part of the factors influencing the proper care of children with asthma. There is currently a wide variety of healthcare provision at the national level. This heterogeneity in access to healthcare providers is illustrated by comparing healthcare provision between different departments and, on a smaller scale, between rural areas and cities. Thanks to the government’s regional observatory website (14), we were able to compare healthcare provision in Martinique with that in the Pyrénées-Atlantiques department.

In terms of demographics, the Pyrénées-Atlantiques department had a population of 706,360 in 2023. There are 787 general practitioners practicing in this area, 56 private pulmonologists, and 37 private pediatricians. In comparison, Martinique had approximately 350,000 inhabitants in 2023. There were 330 private general practitioners. There are 6 private pulmonologists and 7 private pediatricians. Although outpatient consultations are organized by specialists at the University of Martinique Hospital to meet demand (in pediatrics and pulmonology), access to specialist consultations is objectively lower in Martinique.

The provision of general medical care, if we compare the number of general practitioners to the number of inhabitants, is fairly similar (1 general practitioner for every 897 inhabitants in the Pyrénées-Atlantiques, compared to 1 for every 1,060 inhabitants in Martinique).

### 8 - Asthma school

In pediatrics, parents play a fundamental role in the medical care of their children. Among the skills they need to acquire, we will ask them to learn how to recognize an asthma attack, how to recognize the signs of a severe attack, how to administer the necessary emergency treatment by following the action plan, how to use medical devices at home, and how to alert medical personnel at the right time.

By detailing this knowledge, we can understand the importance of education and prevention in ensuring that the child and those around them are prepared in the event of an exacerbation.

In addition, the child must be quickly included in the medical discussion during consultations in order to gradually develop autonomy in their care. All these areas of competence can only be recorded and understood through regular monitoring, consultations focused on prevention, and good coordination between healthcare professionals.

It was precisely in this context that the asthma school was invented in 1992 by the “Asthme et allergie” association in mainland France.

The University of Martinique Hospital has its own asthma school, accredited by the regional health agency since 2015, which provides therapeutic education and a prevention space for families of children with asthma.

It forms an important pillar of care for asthma patients in Martinique through awareness-raising meetings with groups of parents, as well as individualized consultations that allow for a personalized approach to care.

### 9 - Purpose of the study

According to our data, there were approximately 1,600 visits to the pediatric emergency department at the University of Martinique Hospital between January 2023 and December 2023 for asthma attacks. This represents the second most common reason for emergency room visits during this period (after fever), accounting for approximately 7% of visits and requiring hospitalization in 9.6% of cases, representing 154 hospitalizations.

A certain proportion of patients consulting for asthma attacks at the MFME pediatric emergency department did not receive appropriate treatment prior to their arrival, and a significant proportion presented with symptoms classified as mild asthma attacks (according to the PRAM classification). Based on this assumption, we decided to initiate this study.

The main objective of this study is to evaluate the management of asthma attacks prior to consultation at the pediatric emergency department at the University of Martinique Hospital.

Secondly, we will explore data that may explain the poor management of exacerbations in asthmatic children in the pre-hospital setting, by assessing asthma control prior to the attack and describing the medical follow-up of asthmatic patients. The secondary objective is to propose areas for improvement in the medical and paramedical management of asthmatic children in Martinique.

## MATERIALS AND METHODS

### 1 - Study design

We conducted a prospective, descriptive, single-center, cross-sectional study including children residing in Martinique aged between 1 and 18 years old, previously diagnosed with asthma and attending the pediatric emergency department in the university hospital of Martinique due to an asthma exacerbation. We chose a prospective study in order to increase its power, defining in advance the most relevant clinical data to be collected.

Secondly, we examined asthma control in each patient during the month preceding the crisis using the asthma control criteria set out in the GINA guidelines. Several other data were analyzed, such as the patient’s usual medical follow-up, the parents’ feelings about the attack, and the current maintenance treatments. Certain data, such as weight, treatments administered in the emergency department, and parameters on admission to the emergency department, were retrieved retrospectively in order to refine our analyses.

### 2 - Variables

For patient recruitment, questionnaires (Supplementary 1) were distributed in the pediatric emergency department in the university hospital of Martinique to parents consulting for asthma attacks. We took advantage of the long duration of care in the emergency department, due to the frequent need for short-acting bronchodilator inhalation, to distribute the questionnaires to parents accompanying their children, thereby theoretically increasing the response rate among parents. The questionnaire was distributed by the physicians in charge of the asthmatic patient once the clinical evaluation had been completed and treatment had begun.

This questionnaire has four different sections: factors motivating the visit to the emergency room, asthma control criteria, asthma history, parents’ knowledge, and treatments administered at home for the current attack.

The questionnaire takes approximately 15-20 minutes to complete and contains 34 items to check off.

The questionnaires were distributed over a four-month period from July 1 to November 1, 2024.

### 3 - Eligibility criteria

The study focused on evaluating pre-hospital care for asthma attacks in children aged 1 to 18 years, and we included all asthma attacks in patients with known asthma in this age group.

Patients had to meet the clinical and paraclinical criteria for diagnosing an asthma attack, such as dyspnea, wheezing, coughing, and auscultatory wheezing. These attack criteria were not clearly established or imposed on the practitioner. We therefore implicitly accepted each diagnosis once it had been made.

The practitioners who could administer the questionnaires were as follows: senior physicians, residents specializing in general medicine, pediatrics, and emergency medicine working in the pediatric emergency department.

### 4 - Exclusion criteria

We excluded any child presenting their first asthma attack (previously undiagnosed asthma). Patients not residing in Martinique. Patients with a differential diagnosis of dyspnea (such as pneumonia). Patients not accompanied by one of their parents.

We also excluded patients under one year of age (to avoid selection bias related to the bronchiolitis epidemic prevalent during the winter) and patients over 18 years of age. Questionnaires with less than 50% of questions answered were excluded.

### 5 - Data sources

#### 5.1 Crisis treatment

For the therapeutic assessment of asthma attacks in the pre-hospital setting, there are numerous recommendations, and these are not entirely consistent across countries and scientific committees. We referred to the tools used in the pediatric emergency department in the university hospital of Martinique to achieve local and regional consistency in the use of emergency treatments for asthma patients.

We therefore relied on the recommendations of the French university hospital Trousseau in Paris, particularly its emergency action plan, to determine the doses of short-acting bronchodilators to be administered based on the child’s weight. (15) We also used this action plan to determine the time interval between doses of SABA and the timing of oral corticosteroid administration if signs of respiratory distress persisted.

Several criteria had to be taken into account in order to define the quality of therapeutic management of asthma attacks. We therefore developed a score ranging from 0 to 10 in order to assess this as accurately as possible.

○ If SABA is not taken: 0/10

○ If SABA is taken: +3 points

○ So many puffs for its weight: +1 point

○ If oral corticosteroids are taken: +2 points

○ If 3 sets of SABA are taken: +2 points, 2 prises de BDCA: +1, 1 prise de BDCA: +0.

○ If the dosing interval is less than 1 hour: +1 point.

○ If you call the emergency medical services: +1 point.

We therefore estimate a score:

❖ Bad: 0 to 3/10

❖ Moderate: 4 to 6/10.

❖ Good: 7 to 8/10

❖ Perfect: 9 to 10/10.

### 5.2 Asthma control criteria

For the asthma control criterion, we referred to the GINA 2024 recommendations. Asthma control is divided into 3 subgroups based on the symptoms reported by the children during the month preceding the asthma attack.

❖ Controlled (if none of the items are checked).

❖ Partially controlled (if 1 or 2 items are checked).

❖ Uncontrolled (if more than 2 items are checked).

The items in question are as follows: use of SABA more than once a week for children under 6 years of age and more than twice a week for children over 6 years of age; a nighttime cough; asthma-related activity limitations; and daytime symptoms more than twice a week.

This assessment of asthma control will help identify children with poor asthma control who may require an adjustment to their maintenance therapy. Since it has been proven that proper adjustment of maintenance therapy for asthma reduces the risk of asthma exacerbations (in terms of frequency, not severity). (16)

### 5.3 Criteria for optimal medical care

For the criteria for optimal medical care of patients with asthma, we referred to GINA 2024, which defines several criteria for effective disease management:

○ A follow-up visit for asthma within 6 months or 1 year if there are no changes to the treatment regimen, depending on the severity of the asthma. Note that severity is defined by the intensity of treatment required to control the asthma, which is why it is necessary to continually reassess maintenance therapy to determine the minimum effective doses.

○ If a change is made to the maintenance therapy, a follow-up visit must be scheduled in 3 months.

○ Multidisciplinary care is not recommended given the risk that conflicting messages may lead to a disruption in care. Optimal coordination among the various stakeholders is therefore essential.

○ A child with asthma who is over 6 years old is eligible for a pulmonary function testing, which is necessary to assess asthma control in these patients (and thus to adjust their long-term treatment). (17)

○ A pulmonary function test should be performed at least once a year if maintenance therapy is necessary to control asthma. Specialized follow-up care (by a pulmonologist or pediatric pulmonologist) is necessary depending on the severity of the disease (which corresponds to the intensity of the maintenance therapy required to control asthma).

### 6 - Statistical method

The population is described in terms of its count and percentage for qualitative and categorical variables, and in terms of the median and the 25th and 75th percentiles for quantitative variables.

Comparative analyses to calculate the p-value were performed using the Mann-Whitney test for continuous quantitative variables and the chi-square test for binary variables. A p-value less than 0.01 was considered statistically significant.

The size effect was calculated for continuous quantitative variables using a standardized mean difference and for binary variables using the Mahalanobis distance, with a 95% confidence interval calculated.

These analyses, as well as the creation of the figures, were performed using custom Python code (Python 3.9). The p-value was calculated using the scipy package. The effect size was calculated using the effectsize package (https://github.com/nbashir97/effectsize).

### 7 - Ethics

The study was conducted in accordance with French and international guidelines for good practice in epidemiological research. Parental consent was obtained for each questionnaire. Approval from an ethics committee was obtained after the questionnaire and study protocol were presented to the institutional review board (No. 2024/048). The study did not involve any investigations or therapeutic interventions other than those recommended by current guidelines for the management of these patients. Data analysis complies with confidentiality requirements in accordance with the rules of the “Commission Nationale de l’Informatique et des Libertés” (CNIL).

## RESULTS

### 1 - Characteristics of the study population

A total of 183 questionnaires were distributed and collected. Of these, 8 were excluded from the study because the questionnaire was incomplete (<50% response rate). A total of 174 patients were ultimately included in the study.

In this study, 38.3% of the patients were female and 61.7% were male.

Children aged 12–18 are underrepresented compared to the age groups <6 years and 6–11 years (24 patients versus 74 and 76 patients, respectively).

Boys are overrepresented compared to girls, accounting for 61.7% of the patients included. (Table 1)

**Table 1:** Characteristics of the patients included in the study.

|  | Age<6 ans | Age: 6-11 years | Age:12-18 years | Total |
|---|---|---|---|---|
| n = number (%) | 74 (42,3%) | 76 (43.4%) | 24 (13.7%) | 175 |
| Median age (25-75 perc) | 3.5 (2.1-4.7) | 8.6 (7.1 - 10.2) | 14.1 (12.9 - 15.4) | 6.8 (3.9 - 10.2) |
| Female n (%) | 28 (37.8 %) | 31 (40.8 %) | 8 (33.3 %) | 67 (38.3 %) |

### 2 - Pre-hospital care

Patients received a SABA in 88.9% of cases prior to their emergency department visit, and 47% of them received at least three doses of SABA. Among them, the 12- to 18-year-olds had the highest percentage at 62.5%, compared with 42.3% and 45.8% for children under 6 and those aged 6 to 11, respectively. The adjustment of SABA doses based on patient weight varied across age groups: 60.6% of children under 6, 33.8% of those aged 6–11, and 41.7% of those aged 12–18.

The interval between SABA doses is <20 minutes for 54.5% of 12- to 18-year-olds, whereas the percentage of the total study population is 35.2%.

In total, 21.6% of the study population received oral corticosteroids, or slightly more than one in five patients. The 12- to 18-year-olds are the least frequent users, with 16.7% taking oral corticosteroids.

Adolescents seek medical care more quickly than other age groups, with an estimated wait time of <2 hours for 47.6% of them. (Table 2)

**Table 2:** Crisis treatment administered and method of administration prior to the emergency department visit.

| n (%) | <6 years | 6-11 years | 12-18 years | total | p |
|---|---|---|---|---|---|
| N | 74 (42,3%) | 76 (43.4%) | 24 (13.7%) | 175 |  |
| SABA given | 65 / 71 (91.5 %) | 64 / 75 (85.3 %) | 22 / 24 (91.7 %) | 152 / 171 (88.9 %) | 0.439 |
| Inhalations based on body weight | 40 / 66 (60.6 %) | 22/65 (33.8 %) | 10 / 24 (41.7 %) | 72 / 155 (46.5 %) | 0.008 |
| Number of series: |  |  |  |  |  |
| 0 | 6 / 71 (8.5 %) | 11 / 72 (15.3 %) | 2 / 24 (8.3 %) | 19 / 168 (11.3 %) | 0.385 |
| 1 | 16 / 71 (22.5 %) | 7 / 72 (11.1 %) | 4 / 24 (16.7 %) | 28 / 168 (16.7 %) | 0.188 |
| 2 | 19 / 71 (26.8 %) | 20 / 72 (27.8 %) | 3 / 24 (12.5 %) | 42 / 168 (25 %) | 0.301 |
| >=3 | 30 / 71 (42.3 %) | 33 / 72 (45.8 %) | 15 / 24 (62.5 %) | 79 / 168 (47 %) | 0.224 |
| Interval between SABA doses: |  |  |  |  |  |
| <20min | 17 / 58 (29.3 %) | 21 / 61 (34.4 %) | 12 / 22 (54.5 %) | 50 / 142 (35.2 %) | 0.106 |
| 20min-1h | 17 / 58 (29.3 %) | 20 / 61 (32.8 %) | 6 / 22 (27.3 %) | 44 / 142 (31.0 %) | 0.862 |
| 1h-4h | 17 / 58 (29.3 %) | 17 / 61 (27.9 %) | 3 / 22 (13.6 %) | 37 / 142 (26.1 %) | 0.338 |
| >4h | 7 / 58 (12.1 %) | 4 / 61 (6.6 %) | 1 / 22 (4.5 %) | 12 / 142 (8.5 %) | 0.430 |
| Number of breaths in the room >/= 10 breaths or 10 seconds | 29 / 47 (61.7 %) | 29 / 38 (76.3 %) | 5 / 7 (71.4 %) | 63 / 93 (67.7 %) | 0.348 |
| Oral corticosteroids (%) | 16 / 71 (22.5 %) | 17 / 75 (22.7 %) | 4 / 24 (16.7 %) | 37 / 171 (21.6 %) | 0.808 |
| Call to the emergency medical services (%) | 24 / 70 (34.3 %) | 23 / 73 (31.5 %) | 9 / 22 (40.9 %) | 57 / 166 (34.3 %) | 0.714 |
| Time to go to the emergency room n (%): |  |  |  |  |  |
| <2h | 16 / 66 (24.2 %) | 25 / 70 (35.7 %) | 10 / 21 (47.6 %) | 51 / 158 (32.3 %) | 0.102 |
| 2h à 6h | 22 / 66 (33.3 %) | 18 / 70 (25.7 %) | 4 / 21 (19.0 %) | 45 / 158 (28.5 %) | 0.378 |
| 6h à 24h | 21 / 66 (31.8 %) | 13 / 70 (18.6 %) | 5 / 21 (23.8 %) | 39 / 158 (24.7 %) | 0.201 |
| >24h | 5 / 66 (7.6 %) | 6 / 70 (8.6 %) | 2 / 21 (9.5 %) | 13 / 158 (8.2 %) | 0.954 |

SABA: Short-acting β2 agonist

**Figure 1:**
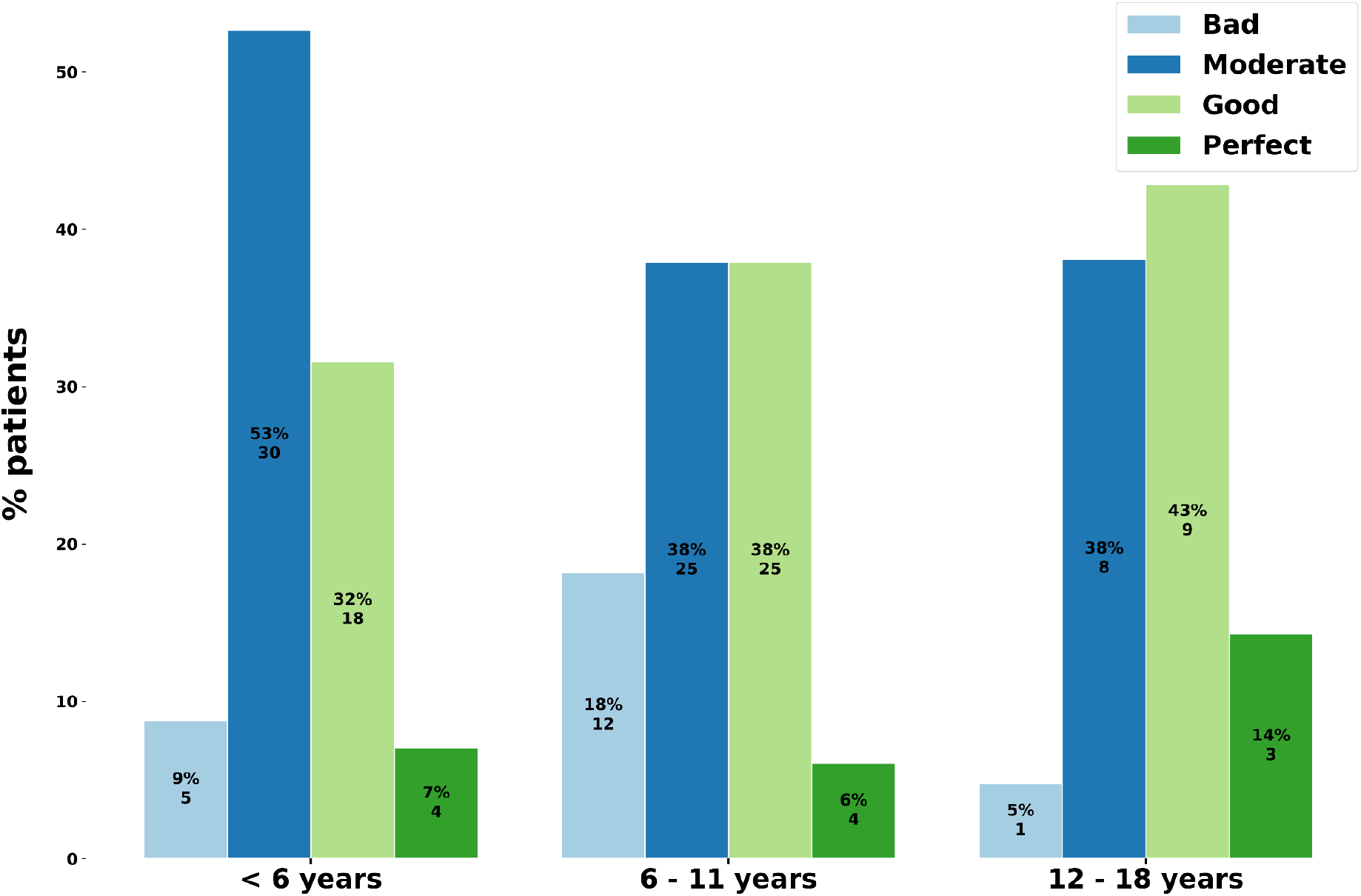
Assessment of prehospital care based on age (score) Figure 1 illustrates the variability in scores by age.

Patients aged 12–18 had the highest scores, with 57% of them achieving a good to perfect score, compared with 39% and 46%, respectively, for the age groups under 6 and 6–11.

More generally, once the scores were applied, the results were as follows (figure 1):

● 11 patients with a perfect score (6.3%)

● 52 patients with a good score (29.3%)

● 63 patients with a moderate score (36%)

● 18 patients with a poor score (10.3%)

● 30 patients not included in the analysis (17.7%)

### 3 - Emergency room care

Upon arrival at the emergency department, the vast majority of patients met the criteria for a moderate asthma attack, accounting for 50.9% of patients with PRAM scores of 4–7. Mild attacks accounted for 32.3% of patients, and severe attacks for 16.8%. Due to a lack of clinical information, 15 patients were not analyzed. (Table 3)

**Table 3:** PRAM upon arrival at the emergency department.

| n(%) | Total |
|---|---|
| PRAM 1-3 | 52 /161 (32.3%) |
| PRAM 4-7 | 82 /161 (50.9%) |
| PRAM 8-12 | 27 /161 (16.8%) |

The median oxygen saturation upon arrival at the emergency department was 96%.

The vast majority of patients received corticosteroid therapy (oral or IV), accounting for 76.7% of patients. The median number of nebulizer treatments administered was 3, with little variation across age groups.

Among the patients in the study, 18.8% required stabilization and therapeutic management in the resuscitation unit. Patients under 6 years of age were underrepresented, with 12.5% transferred to the resuscitation unit, compared to 22.4% and 25% for those aged 6–11 and 12–18, respectively.

14.6% required hospitalization following their visit to the emergency department; 85.4% were discharged home.

The median time spent in the emergency department for all age groups is 322 minutes, or approximately 5 hours and 20 minutes. However, there is significant variability in the time spent in the emergency department, as illustrated by the standard deviation between the 25th and 75th percentiles (192–833 minutes).

### 4 - Asthma control criteria in the month prior to the attack

The results indicate that 35.4% of children had well-controlled asthma in the month prior to the crisis, 33.1% had partially controlled asthma, and 24% had uncontrolled asthma. Children aged 6–11 had the highest percentage of well-controlled asthma. Among those aged 12–18, 37% had poorly controlled asthma one month before the crisis.

Among patients with uncontrolled and partially controlled asthma, 53.3% had seen their doctor within the past six months. (figure 2)

**Figure 2:**
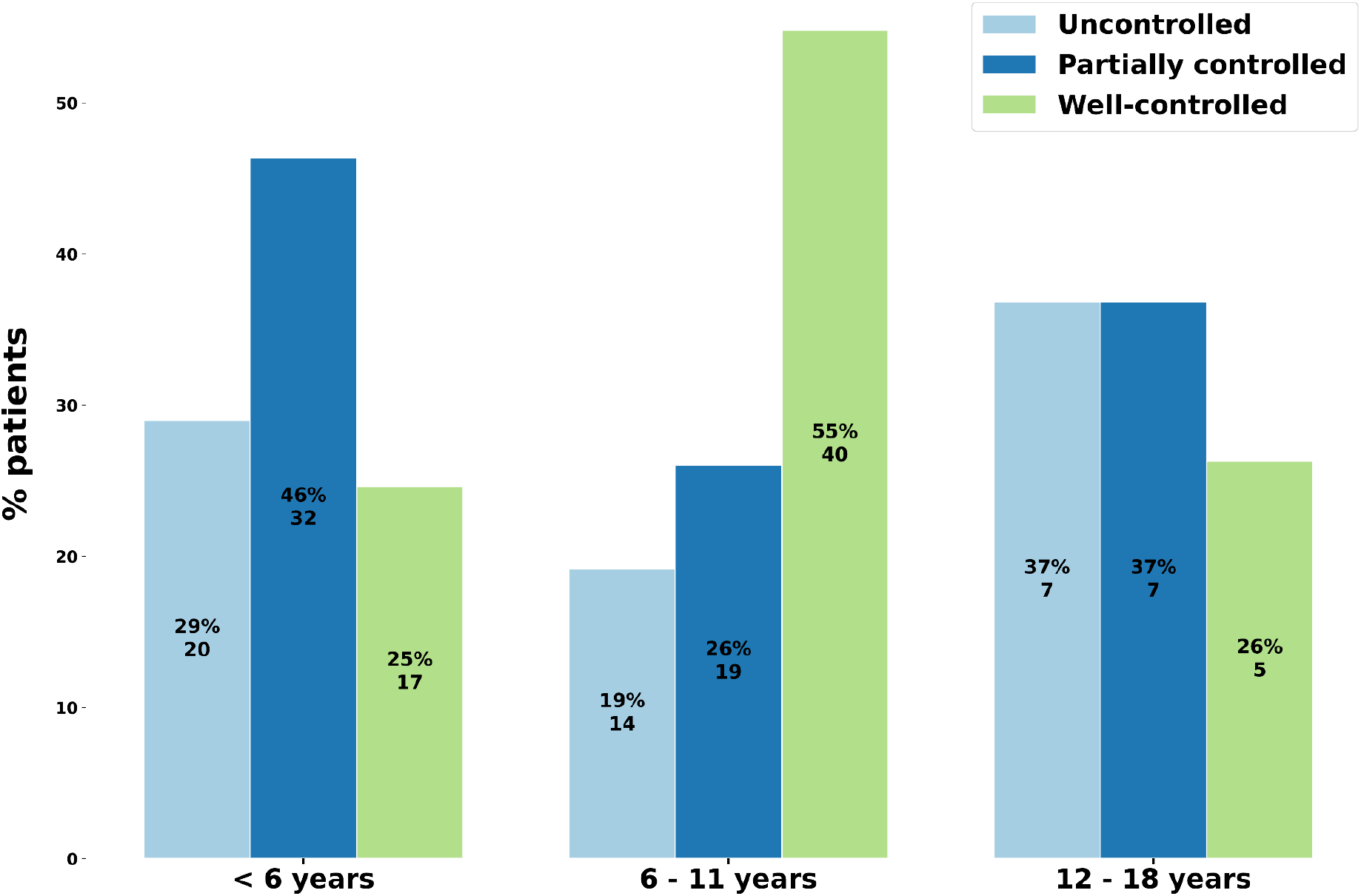
Assessment of asthma control prior to the attack. Proportion of patients with well-controlled, partially controlled, or uncontrolled asthma by age.

Among the well-controlled patients, 16 out of 62 (25.8%) were treated in the resuscitation unit, 11 out of 61 (18%) were admitted to a general ward for their asthma attack, and none were transferred to the intensive care unit. Among the partially controlled patients, 8 out of 56 (14.3%) were treated in the resuscitation unit, 8 out of 56 (14.3%) were admitted to the hospital, and 1 out of 56 (1.8%) was transferred to the ICU. Among the uncontrolled patients, 3 out of 38 (12.8%) were treated in the resuscitation unit, 4 out of 39 (10.3%) were admitted to a general ward, and 2 out of 39 (5.1%) were transferred to the ICU. (supplementary figure 1).

### 5 - Medical care for children

Among 6- to 11-year-olds, care by a pulmonologist is more common than in other age groups, accounting for 12% of patients. Pediatric care appears to be most common in early childhood, with 14% of children under 6 years of age receiving it, compared to 5% and 0% of children aged 6–11 and 12–18, respectively.

Care is primarily provided by a general practitioner for children under 6 and those aged 6–11 (43% and 50%, respectively), whereas it accounts for 30% of care for those aged 12–18.

Furthermore, among 12- to 18-year-olds, 30% of care is provided by a general practitioner, 30% by an allergist, and 30% receive no care for their asthma. (figure 3)

**Figure 3:**
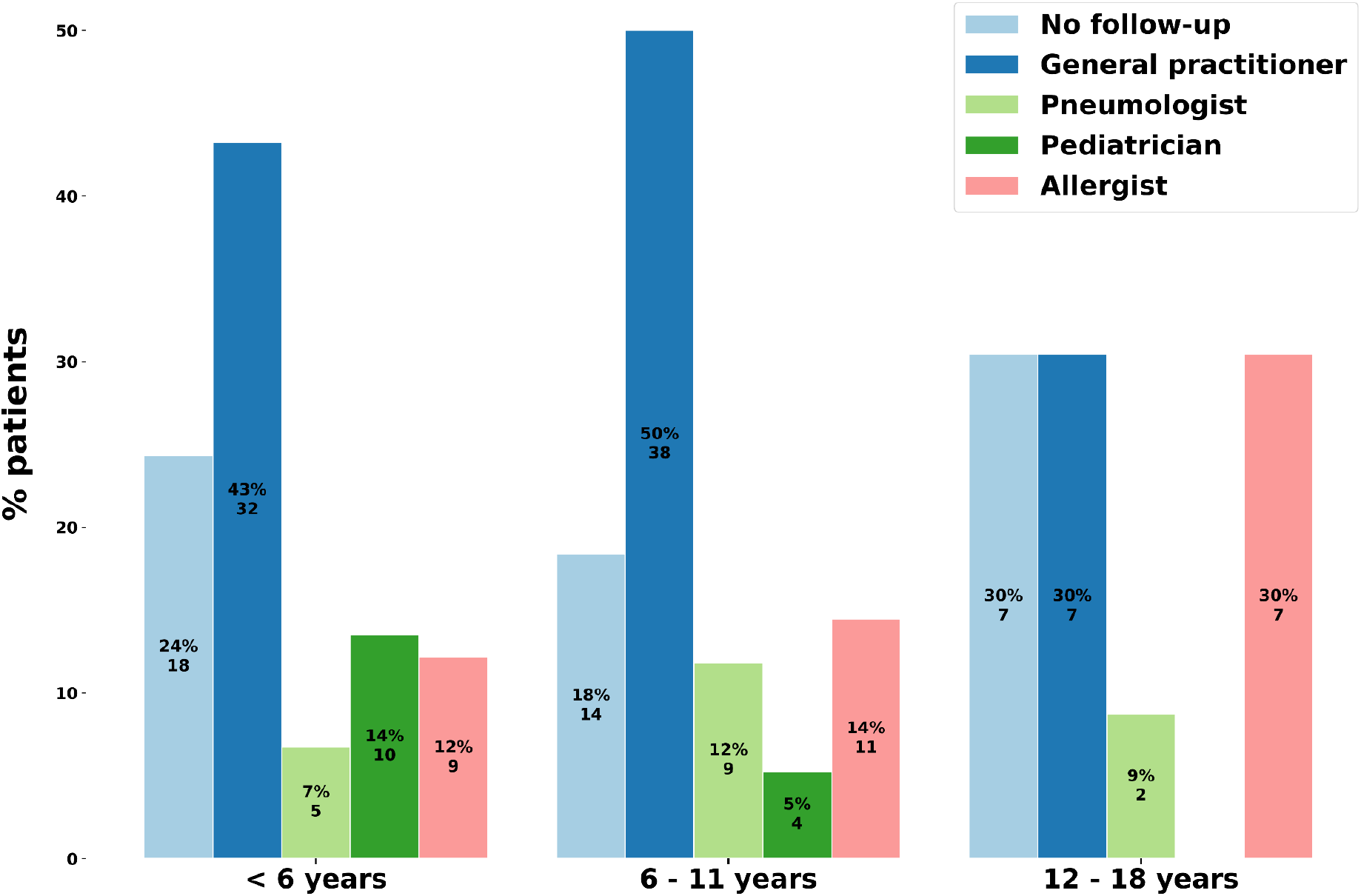
Characteristics of long-term medical care for patients with asthma.

Children aged 11–18 represent the population with the highest percentage of pulmonary function tests performed, at 35%, compared with 8.6% among children aged 6–11. Long-term asthma treatment was reported in 46.8% of the total sample.

A higher proportion of children aged 1–6 had an asthma action plan—62% of them—compared with 44.7% of those aged 6–11 and 45.5% of those aged 11–18. When an action plan was in place, an average of 89.8% of patients reported having used it.

Asthma school was known to 42.9% of children aged 11–18, compared with 7% and 15.8% of children under 6 and those aged 6–11, respectively. (table 5)

In total, 64% of patients receiving maintenance therapy were treated with Flixotide, while 20.4% were treated with Seretide; only 2 patients were on a ICS/formoterol combination, and 1 was on a ICS/vilanterol combination

An individualized action plan was developed for 65.9% of patients, and 51.8% of patients had a home action plan.

### 6 - Personal experiences of the asthma attack

In our study, 82% of parents reported feeling that their child was in danger before going to the emergency room. Only 49.1% reported feeling a loss of control in the face of the crisis, and 86.7% reported having used all the resources available to them before seeking care at the pediatric emergency department.

## DISCUSSION

This study was conducted as a cross-sectional, prospective study; the response rate to the questionnaire was good, allowing for the enrollment of 175 patients out of the 181 questionnaires distributed during the period from July through the end of October 2024.

The primary objective of the study was to describe and evaluate the management of asthma attacks in children aged 1 to 18 years prior to presentation at the pediatric emergency department. One of the reasons for conducting this study is the high prevalence of asthma in Martinique, as well as the significant challenges in managing this chronic disease, due to the variety of factors required for its optimal clinical control.

### 1 - Characteristics of the study population

The vast majority of patients included in the study were between 1 and 11 years old, accounting for 86.3% of the study population.

Adolescents aged 12 to 18 accounted for 13.7% of the study population.

Adolescents are a difficult-to-treat population; they rarely seek medical care and tend to present to the emergency department only when their asthma has actually deteriorated significantly, with severe asthma attack criteria, thereby increasing the risk of serious respiratory decompensation and admission to the intensive care unit.(18)

Boys are overrepresented compared to girls, accounting for 61.7% of the completed questionnaires. These figures are not surprising, since girls typically develop asthma symptoms primarily during adolescence.(19)

### 2 - Analysis of the results

#### 2.1 Analysis of the primary criterion

Several factors come into play in the proper management of an asthma attack—both from a therapeutic standpoint, involving various treatments, and from a behavioral standpoint, particularly the ability to recognize signs of respiratory distress, the ability to alert others, and the appropriate waiting time before seeking emergency care.

This multifactorial nature has made it difficult to accurately assess the management of asthma attacks in the prehospital setting. To evaluate it comprehensively, we developed a score ranging from 0 to 10 that combines several management criteria we consider important.

To develop this score, we ranked the priority interventions for the proper management of asthma attacks in children.

We felt that the most important intervention was SABA treatment at home (3 points). The results are detailed in Table 2. Slightly more than 1 in 10 patients (11.1%) did not receive a SABA before their emergency department visit. Among the 88.9% who took SABA, 52.5% administered at least 3 doses. (2 points)

Furthermore, 68% of patients had a valid prescription for Ventolin or Ventolin at home. This finding highlights the limitations of attributing poor prehospital therapeutic management solely to parents; a lack of follow-up and foresight can affect asthma attacks. Management of an asthma attack involves two distinct approaches depending on the severity and tolerance of the attack. If symptoms are mild, it may be recommended to space out SABA doses by 4 hours and repeat as needed until symptoms improve. If the attack worsens or is moderate or severe from the outset, administering SABA every 20 minutes for 1 hour is indicated. The questionnaire did not distinguish between these two situations and could therefore be confusing. We decided to award only one point for this parameter, and 66.2% of patients maintained an interval of <1 hour between Ventolin doses.

The instruction to call the emergency medical services is included in the action plan provided by the emergency department. However, only 34.3% called the emergency medical services before going to the emergency department. Interestingly, 72.7% of our patients report having received their crisis action plan from the pediatric emergency department and should therefore, in theory, call the SAMU if the crisis persists and they detect signs of a serious condition. (1 point)

Only 21.6% of patients had taken oral corticosteroids before their emergency department visit. (2 points) One might also question the availability of these medications at home, particularly when the last attack occurred some time ago. (lost prescription, expired medication, no preventive follow-up visit).

SABA doses are tailored to the patient’s weight for 62.5% of children under 6 years of age, 33.8% of children aged 6–11, and 41.7% of adolescents. This difference can be explained by the fact that the minimum doses are close to the recommended doses for children under 6 years of age with low body weight. Furthermore, physicians’ recommendations and practices vary by approximately 2 puffs depending on the child’s weight. Overall, 47.1% of patients received a SABA dose appropriate for their weight. We chose not to include the time elapsed before seeking emergency care in the score, since some patients reported noticing the first signs the day before and then observing an unfavorable progression (thus estimating a delay of >24 hours even though their response to the crisis was appropriate).

Figure 1 shows a moderate difference in scores between children under 6 and those over 6. This finding could be explained by several factors, including the recent discovery of the disease and parents’ limited knowledge of it. Special attention could be given to the under-6 age group, particularly in strengthening prevention measures.

There is currently significant room for improvement in the prehospital management of asthma attacks. Focusing our efforts on this phase of asthma could reduce the morbidity associated with the disease. Ultimately, we could significantly reduce the psychological impact on patients and improve their quality of life by decreasing the number of emergency department visits, as well as by reducing anxiety related to asthma attacks through a better understanding of the disease. One of the goals is to reduce the social impact of the disease, such as school absenteeism.

The number of visits to pediatric emergency departments could decrease, which would improve the quality of care for other patients by reducing the workload of healthcare providers.

Healthcare costs related to asthma are currently substantial, and a shift toward prevention-focused care could not only improve the quality of patient care but also reduce public spending in the long term.(20)

All of these factors must be taken into account to improve asthma care in Martinique.

### 2.2 Analysis of secondary objectives

#### 2.2.1 Asthma control

To assess asthma control, we relied—as described in the methods section—on the GINA 2024 recommendations.

We found that 35.4% of the children were well-controlled in the month prior to the crisis. 33.1% were partially controlled, and 24% were uncontrolled.

“Among uncontrolled and partially controlled patients, 53.3% saw their doctor within 6 months” (Figure 2). This estimate is likely an overestimate, given that the study period included the start of the school year. It is possible that many of the reported follow-up visits were solely for the purpose of completing the individualized welcome plan required by elementary and middle schools. Slightly fewer than one in two patients therefore showed signs of poor asthma control in the previous month; these warning signs—such as a loss of control over asthma symptoms—should, in theory, alert the family and lead to a medical consultation for reevaluation.

One of the factors complicating this analysis is the inclusion of patients currently under follow-up for poorly controlled asthma who may be in the therapeutic adjustment phase.

We investigated whether there might be a link between the degree of asthma control and the potential severity of an asthma attack in our population.

We observed that the number of patients treated in the emergency resuscitation unit was higher among well-controlled patients, but that the number of children transferred to the intensive care unit was higher among partially or poorly controlled patients (3 in total). Given the small proportion of patients requiring intensive care (3 out of 174), it is difficult to draw conclusions about a causal relationship.

Some studies have found a causal link between poor clinical control and morbidity and mortality associated with asthma attacks. In particular, by assessing the excessive use of SABAs over a given period—an indirect marker of asthma control—the results show a higher risk of asthma exacerbations and asthma-related mortality in this population.(21) (22) Furthermore, anyone with asthma is at risk of a severe attack, which is why monitoring and prevention are so important. Even in patients with few symptoms—who haven’t had an asthma attack in years—it’s essential to have the tools to deal with one if it occurs.

In 2009, a French study highlighted the low proportion of patients with controlled asthma among those seeking care at the emergency room. (23) In this study, conducted in primary care settings, 3,431 patients were included; of these, 26% had good asthma control, 41.3% had moderate control, and 32.7% had poor asthma control. This is an older but compelling study, with a different research approach (recruitment from primary care settings). Despite this, the percentage of patients with well-controlled asthma in our study is higher. Although 13 years have passed since that study, and setting aside any differences and biases, we can assume that asthma control is slightly better today in Martinique than it was in mainland France in 2009.

#### 2.2.2 Medical management of asthma

According to current GINA recommendations, a child with asthma should be monitored by a primary care physician without the need for a specialist consultation if the asthma is well-controlled and of mild severity. It should be noted that the definition of asthma severity depends on the intensity of the maintenance therapy prescribed to control symptoms. Referral to a specialist should therefore be considered if asthma control proves difficult and requires an increase in dosage or a change in the type of maintenance therapy, classifying the asthma as moderate to severe.

In our study, 22.3% of parents reported that their child’s asthma was not being medically managed. This represents nearly 1 in 5 children who visit the emergency room. Among children receiving medical care, 54.8% of those under 6 years of age are managed by their primary care physician, and 16.7% of them are seen by a pediatrician. The transition from pediatric care to primary care appears to occur around age 6, since 70.3% of children over 6 years of age receive asthma care from their general practitioner.

Medical management of asthma by an allergist is around 25% among patients over 6 years of age, compared to nearly 12% managed by a pulmonologist. There may be a bias in these data, since one pulmonologist (out of the six private-practice pulmonologists in Martinique) specializes in allergology.

As described in the introduction, the availability of care in Martinique is limited in the private-practice specialties of pulmonology, allergology, and pediatrics. General practitioners therefore play a central role in the care of patients with asthma, regardless of their age.

In terms of follow-up frequency, 51.5% of patients reported having had a follow-up visit for asthma within the previous 6 months, and 70% within the past year.

Medical recommendations for the frequency of follow-up visits for asthma are as follows: 3 months after starting a new treatment or making a recent change in therapy, then every 6 to 12 months depending on the severity and control of the asthma.

Our study does not examine the quality of follow-up visits or the topics discussed during these visits—particularly those focused on prevention and patient education. This could be an area for evaluation and improvement in the future.

One of the most notable figures in this study is the percentage of children with asthma who are on maintenance therapy—only 46.8% (Table 5). The proportions vary little across the different age groups.

The range of treatments used in this study is limited, even though many treatment options are available. This may be due to prescribing practices. One limitation of our assessment is the lack of data on the exact doses administered (which could reveal cases of treatment failure and indications for treatment changes).

To date, studies have shown the importance of long-term treatment with ICS in mild and moderate asthma for reducing the risk of asthma attacks (24), a reduction in morbidity and mortality and an improvement in pulmonary respiratory function in adulthood. (12)

One potential barrier to their use is the known side effects of ICSs. However, at low to moderate doses, the most commonly reported side effects are mild and preventable: oral thrush is the most common and can sometimes lead to dysphagia and coughing. One of the known adverse effects of corticosteroids in children is reduced height growth; however, low- or moderate-dose ICSs are associated with an average loss of 1% in height growth. (25) Central effects (Cushing’s syndrome, osteoporosis, cataracts, etc.) are very rare with ICS.

Compared to other studies, we achieved good rates of action plan implementation, although there is still significant room for improvement. (26)

One of the notable figures is the percentage of patients who reported receiving an action plan in the pediatric emergency department. This reflects the consistent practice of providing action plans upon discharge from the pediatric emergency department at the MFME.

Pulmonary function testing was performed in only 14.3% of children over the age of 6. Yet this is a key laboratory test for the diagnosis and follow-up of patients with asthma. Several possible explanations can be considered: lack of access, lack of training, and lack of data (particularly regarding its practical application and usefulness for adjusting treatment). In patients with mild clinical symptoms or whose last asthma attack occurred some time ago, pulmonary function testing could be useful for reassessing the treatment plan and improving patient adherence. It should be noted that none of the 17 patients admitted for conventional hospitalization had undergone pulmonary function testing in the past.

Peak expiratory flow is rarely used; only 4.1% of patients over the age of 6 had one at home, and 13.4% believed they knew how to use it. Yet it can be a useful and effective tool both during acute episodes and for chronic monitoring.

Although there appear to be improvements in the management and treatment of asthma in France compared with studies conducted in France and Europe over the past 15 years, our study confirms that inadequate treatment and a lack of follow-up for asthma remain ongoing issues. (27)

#### 2.2.3 Personal experience with an asthma attack

Parents’ experiences are an important factor to consider. A qualitative study showed that parents still had significant concerns about the side effects of asthma treatments, and that these concerns could interfere with the administration of treatments at home. (28)

If we objectively compare the estimate that all available resources were exhausted with the proportion of patients who received at least three doses of SABA (82 out of 174, or 47.1%) or the number of patients who received oral corticosteroids before arriving at the emergency department (21.6%); a gap becomes evident between parents’ perceptions and the actual therapeutic management of asthma attacks.

Parents’ anxiety during asthma attacks may reflect a lack of experience, information, and sometimes understanding of the disease. It would be interesting to study parents’ fears and concerns in Martinique and to adapt medical and paramedical communication accordingly.

Furthermore, only 27.6% of patients in the study consulted a primary care physician for an asthma attack within 5 days prior to their emergency department visit. It can therefore be estimated that the vast majority of treatments administered during asthma attacks in this study were primarily based on parents’ knowledge and habits.

The morbidity associated with asthma is significant among both children and their caregivers. It would be worthwhile to assess this morbidity in Martinique using questionnaires such as the PAQLQ and to raise patients’ awareness of the concept of quality of life. A prospective study conducted in 2019 in Algeria demonstrated a correlation between the degree of poor asthma control and the level of morbidity associated with asthma in both children and their parents. (9) The repercussions for children are social (school absenteeism, reduced participation in sports) as well as psychological (isolation, stigmatization) and can adversely affect their overall health.

#### 2.2.4 Medical care in the pediatric emergency department

Table 3 shows that 32.3% of patients had a PRAM score between 0 and 3 upon arrival at the emergency department. This means that nearly 1 in 3 children presenting to the emergency department had symptoms of a mild asthma attack. These figures highlight the need for therapeutic education for parents on understanding and recognizing the signs of a severe asthma attack. This aspect of prevention is particularly challenging to address given the emotional component, parents’ lack of theoretical knowledge, and their lack of experience with asthma attacks. These figures do not take the patients’ medical history into account. Patients may have exhausted all available treatments and, due to the persistence of their symptoms, sought care in the emergency department.

One of the interesting variables in table 4 is the time spent in the emergency room, which averages 5 hours and 20 minutes across all age groups. Since exacerbations often occur at night, one can imagine the impact an episode can have on the following days, and the resulting decline in quality of life for patients whose condition is not well controlled.

**Table 4:** Characteristics of emergency department care.

| n (%) | <6 years | 6-11 years | 12-18 years | total |
|---|---|---|---|---|
| Median O2 saturation on admission (25th–75th perc) | 97% (95 - 97.8) | 96% (94 - 98) | 96 (94 - 99) | 96% (95 - 98) |
| Median number of SABA nebulizations in the emergency department (25th–75th perc) | 3 (3 - 6) | 3.5 (2.8 - 8) | 3 (2.5 - 6.2) | 3 (3 - 10) |
| Corticosteroid therapy in the emergency department n(%) | 57 (79.2%) | 60 (77.6%) | 16 (66.7 %) | 133/172 (76.7 %) |
| Resuscitation unit n (%) | 9 (12.5%) | 17 (22.4 %) | 6 (25.0 %) | 32 / 172 (18.8 %) |
| Hospitalizations n (%) | 8/ 71 (11.3 %) | 15 (19.7 %) | 2 (8.3 %) | 25 / 171 (14.6 %) |
| Median time spent in the emergency department in minutes (25th–75th perc) | 324 (184 - 481) | 350 (203- 1346) | 250 (180 - 690) | 322 (192 - 833) |

perc: percentile

**Table 5:** Characteristics of asthma patients seen in the pediatric emergency department, by age.

| n (%) | 1 à 6 years | 6 à 11 years | 11 à 18 years | total | p |
|---|---|---|---|---|---|
| **Long-term treatment :** |  |  |  |  |  |
| present: | 30/65 (46.1%) | 31/71 (43.7%) | 10/21 (47.6%) | 74/158 (46.8%) | 0.819 |
| absent: | 37 / 66 (56.1 %) | 36 / 71 (50.7 %) | 11 / 21 (52.4 %) | 84/159 (52.8 %) | 0.491 |
| unknown: | 4 / 70 (5.7 %) | 5 / 76 (6.6 %) | 0 / 21 (0.0 %) | 9/168 (5.4 %) |  |
| **Individualized Action Plan:** | 43/72 (59.7 %) | 52/73 (71.2 %) | 14/21 (66.7 %) | 110/167 (65.9 %) | 0.343 |
| **Action plan at home:** |  |  |  |  |  |
| present | 44 / 71 (62.0 %) | 34 / 76 (44.7 %) | 10 / 22 (45.5 %) | 88/170 (51.8 %) | 0.090 |
| used if present | 40 / 44 (90.9 %) | 31 / 34 (91.2 %) | 8 / 10 (80.0 %) | 79 / 88 (89.8 %) | 0.556 |
| delivered at the emergency | 29 / 44 (65.9 %) | 27 / 34 (79.4 %) | 8 / 10 (80.0 %) | 64 / 88 (72.7 %) | 0.356 |
| **Valid prescription for Ventolin for home use: :** | 47 / 71 (66.2 %) | 53 / 75 (70.7 %) | 14 / 22 (63.6 %) | 115/169(68.0%) | 0.763 |
| **Asthma school:** |  |  |  |  |  |
| known and visited | 5 / 71 (7.0 %) | 12 / 76 (15.8 %) | 9 / 22 (40.9 %) | 26/170 (15.3 %) | <0.01 |
| unknown uniquement | 20 / 71 (28.2 %) | 16 / 76 (21.1 %) | 3 / 22 (13.6 %) | 39/ 170 (22.9 %) | 0.314 |
| unknown | 46 / 71 (64.8 %) | 48 / 76 (63.2 %) | 10 / 22 (45.5 %) | 105/170 (61.8 %) | 0.245 |
| **Last follow-up visit for asthma:** |  |  |  |  |  |
| 0 - 6 months | 35 / 69 (50.7 %) | 40 / 72 (55.6 %) | 9 / 21 (42.9 %) | 84/ 163 (51.5 %) | 0.574 |
| 6 months - 1 year | 14 / 68 (20.6 %) | 12 / 71 (16.9 %) | 4 / 21 (19.0 %) | 30/ 161 (18.6 %) | 0.856 |
| > 1 year | 11 / 68 (16.2 %) | 18 / 71 (25.4 %) | 7 / 20 (35.0 %) | 37 / 160 (23.1 %) | 0.160 |
| **PFT performed** |  | 6 / 70 (8.6 %) | 7 / 20 (35.0 %) | 13 / 91 (14.3 %) |  |

PFT: pulmonary function test (for patients > 6 years of age)

76.7% of patients receive oral or systemic corticosteroid therapy, while 67.7% of patients present with a moderate-to-severe attack upon arrival. Treatment with oral corticosteroids is not indicated for mild attacks (PRAM 0–3). These figures may suggest that treatment in the emergency department is sometimes excessive, but they may also reflect the rapid clinical worsening of attacks in some patients. This interindividual variability can undermine standardized care protocols and requires additional vigilance on the part of medical staff.

### 2.3 Comparison with the literature

A fairly similar study was conducted in France at the pediatric emergency department in Clermont-Ferrand between November 2010 and February 2011. (26) This is a prospective, descriptive study that aims to evaluate prehospital care for children with asthma and to propose ways to improve the management of asthma attacks. In this study, 55.9% of patients used SABA before arriving at the emergency department. 27.3% had been prescribed oral corticosteroids.

They estimated that 17.5% of patients received appropriate prehospital care (based solely on the administration of SABA and oral corticosteroids). Among the asthma patients in this study, 48.8% were on maintenance therapy. 45.2% of the children were not receiving follow-up care for asthma, and 32.2% were being followed by a pediatrician or pulmonologist. A written action plan had already been provided to families in 16.4% of cases. Only 22.7% had an individualized care plan.

In our study, 88.9% of patients used SABA before visiting the emergency department. However, 52% took at least 3 doses of SABA. The use of SABA therefore appears to be fairly similar. The focus for improvement seems to lie in administration methods (number of puffs, dosing intervals). An assessment of inhalation techniques could not be conducted during this study and is a priority, particularly to limit adverse effects. (A simple improvement in techniques could lead to a reduction in prescribed doses.)

In our study, only 21.6% of patients had used oral corticosteroids before arriving at the emergency department. This represents a significant difference. It might be worthwhile to explore the barriers to parents administering oral corticosteroid treatments in an emergency setting (access to treatment, feared side effects, beliefs, etc.).

### 3 - Limitations of the study

This is a single-center study. To conduct this study, we chose to examine a population of patients presenting to the pediatric emergency department for an asthma attack. Assessing the primary outcome measure required prioritizing the importance of treatments and emergency interventions using a scoring system. This score is entirely subjective (since we developed it ourselves), but it allows for a more comprehensive understanding of hospital care (rather than focusing solely on the administration of oral corticosteroids and SABA, as seen in other studies). The heterogeneity of (mild) recommendations—but especially of medical practices—introduces a bias in the evaluation of prehospital care for patients. A 2015 doctoral thesis conducted in Martinique illustrated this diversity in asthma management in primary care. The study’s assessment of prehospital care is therefore naturally influenced not only by patients’ understanding but also by medical practices that are sometimes inconsistent.

This inconsistency in the messages conveyed to patients can sometimes reduce adherence to or understanding of medical advice, especially among patients receiving multidisciplinary care. Standardizing medical advice is a priority not only to facilitate the management of asthma attacks but also to control asthma in children.

By choosing to recruit patients in the emergency department, one may be exposed to a population of children with asthma who are predominantly poorly controlled or poorly managed medically, and may underestimate the number of well-controlled and well-managed patients in the general population. Indeed, a close link between poor asthma control and the risk of asthma exacerbations is well known and has been described. (29) A follow-up study conducted in primary care could, in the future, supplement our data and help identify specific areas for improvement in patient care.

Our study did not examine the predominant socioeconomic backgrounds of emergency department patients or the impact of these backgrounds on the use of healthcare services. This area of study is particularly relevant in Martinique, since 32% of Martinique’s children live below the poverty line—twice the rate in metropolitan France. We chose not to investigate social vulnerability, as we felt that the questionnaire format and the setting in which it was completed were not conducive to a proper assessment. (We could have examined educational attainment, parents’ occupations, or single-parent households, for example).

Our study briefly touched on how families feel during an asthma attack. This research topic is, however, of interest, particularly for understanding the emotional impact on the quality of home-based therapeutic care. Strategies to help manage anxiety related to asthma attacks could be developed accordingly. The assessment of environmental risk factors and the identification of factors that trigger asthma attacks were also not investigated.

These decisions were made with the aim of encouraging parents to complete the questionnaires in full, given the time constraints and the fact that the environment was sometimes not conducive to doing so.

### 4 - Areas for improvement

#### 4.1 Primary Care

Asthma is a highly prevalent disease affecting populations worldwide. There are numerous studies and medical articles on the subject, which have enabled the gradual evolution of medical recommendations aimed at improving the management of asthma. Unfortunately, changes in expert recommendations can sometimes occur more rapidly than changes in medical practice. (30). This phenomenon has been observed in conditions other than asthma, notably bronchiolitis and the treatment of nasopharyngitis. (31) Differences in care and a lack of updated knowledge can affect all medical specialties, particularly within a single emergency department. (32)

Regular continuing education for physicians and the standardization of care based on guidelines could ultimately improve asthma management in Martinique. Several types of continuing education tools aimed at healthcare providers (residents, general practitioners, and specialized nurses) can be—and are being—developed in the form of seminars and online training courses, for example.

Standardizing practices requires the use of the same guidelines and the same tools. A collaborative effort could be undertaken with the adult pulmonology department, pediatric pulmonologists, referring general practitioners, and specialized nurses to develop uniform action plans and individualized care plans throughout Martinique. And perhaps a digital platform could be created to make these documents available.

A computer-based tool to assist physicians in decision-making—particularly regarding the initiation or adjustment of treatment—could also be developed.

Increasing the recruitment of physicians specializing in pulmonology and pediatric pulmonology could lighten the already very diverse workload of general practitioners and help ensure better patient care. To achieve this, the region must be attractive to these specialists; however, raising awareness of the challenges involved in managing asthma and highlighting current initiatives to develop and improve the healthcare network could potentially attract more than a few of them.

Helping to develop primary care clinics—which serve as a screening mechanism before patients seek emergency care—and establishing common, standardized protocols with emergency services in the event of a crisis could be another area of focus.

The lack of reimbursement for therapeutic education consultations—which are generally lengthy—does not encourage preventive care in private practice. A specific fee schedule tailored to therapeutic education consultations could be one of the tools used to change practices in private practice.

#### 4.2 Emergency medicine

Before visiting the emergency room, 34.3% of patients contacted emergency medical services. A significant proportion of children seen in the emergency department have mild episodes that can be treated at home. It is possible that we underestimated this patient population in our study, since the duration of visits to the emergency department is short and does not allow sufficient time to complete the questionnaire. One area for improvement could be to increase the number of emergency medical services staff and perhaps create a specialized pediatric dispatch unit. (33)

In the emergency department, the top priority is treating the child’s asthma attack. But once treatment has been initiated and the attack has subsided, the priority shifts to preventing a relapse in the coming days. To minimize the chances of recurrence as much as possible, trained medical or paramedical staff must conduct a prevention session to address key topics such as the use of inhalers, the importance of ongoing treatment, the avoidance of environmental risk factors such as tobacco, and the need for close medical follow-up after the child returns home (re-evaluation within 3 days). This information must be provided by trained medical or paramedical personnel.

Part of the medical team could be specifically trained in post-crisis preventive measures (for example, an advanced practice nurse or a nurse trained in asthma care within the team, and a referring physician on the medical team). (34)

The initiation of maintenance therapy in the emergency department is an important issue, as many clinicians do not consider the setting and timing to be appropriate. However, we found no studies on this topic. The risk of patients losing vision after their visit to the emergency department and the beneficial effect of ICS on the risk of asthma exacerbations suggest that emergency physicians should prescribe maintenance therapy, in conjunction with close follow-up care.

In our study, 14.8% were admitted to a general ward, while 29.7% of patients received more than 6 SABA nebulizer treatments.

According to the SP2A’s new guidelines, hospitalization is recommended for children with a PRAM greater than 4 despite 6 rounds of nebulization. It appears that the hospitalization rate is too low compared to what is recommended. If the general ward’s capacity allowed it, hospitalization could serve as an anchor point in the initiation or follow-up of asthma care. A direct link with healthcare providers could be established. Access to pulmonary function testing, a connection to asthma education programs, the initiation of specialized care if necessary, and a focus on prevention and education for parents and children. Psychological support could be offered if necessary.

Often, there are few hospital beds relative to the needs arising in the emergency department, and decisions must be made based on the theoretical benefit of hospitalization for the patient. A study on the use of peak expiratory flow in the emergency department could be an interesting avenue to explore, particularly among children for whom the decision to admit or discharge is under discussion.

Given the shortage of inpatient beds and the fact that asthma attacks often improve over time, one option would be to establish a post-emergency care clinic to be visited three days after discharge from the emergency department.

An appointment would then be scheduled in the emergency department, and an automated message could be sent to families to remind them of the appointment date and time, thereby encouraging them to attend the appointment.

### 4.3 Therapies and tools

#### 4.3.1 Raise awareness about the completion of pulmonary function testing

Pulmonary function testing is the cornerstone of paraclinical evaluations in children with asthma. It allows for an accurate assessment of the patient’s lung function and is useful for monitoring, adjusting treatment, and evaluating the impact on respiratory function over time.

Given the clinical heterogeneity of asthma, its use can accurately assess a patient’s respiratory function and guide the clinician in managing the patient’s care.

In our study, 14.3% of children over the age of 6 had access to pulmonary function testing.

A study could be proposed among primary care physicians to assess their attitudes toward the use of pulmonary function testing, as well as their self-assessment of their ability to interpret the results and the barriers to prescribing it (difficulties in access, lack of familiarity, and lack of training).

#### 4.3.2 Using the peak flow meter

In our study, only 4 children had a peak flow meter at home, and 13 caregivers believed they knew how to use it.

The EDELMA study, conducted in 2018 at the CHUM’s asthma school, found that 4 of the 108 children included in the study reported that they had mastered the device.

To date, the use of peak flow measurement is recommended primarily for patients with mild asthma symptoms or those who are unable to assess the severity of their attack. This tool provides a visual assessment of the breathing difficulty associated with the attack and may allow for faster initiation of treatment, thereby potentially increasing the effectiveness of emergency therapy.

A prospective study conducted in 2024 demonstrates the usefulness of peak flow measurement in assessing how children with asthma perceive their respiratory limitations. Furthermore, after more than 6 months of use, it appears to be beneficial for treatment adherence (consistent use of inhaled corticosteroids) and changes in respiratory function (improvement in peak expiratory flow). (35)

Selon le GINA 2024, le peak flow meter a sa place pour l’estimation de la fonction respiratoire chez les patients n’ayant pas accès à la spirométrie. Une adaptation thérapeutique à l’aide du débit expiratoire de pointe serait plus recommandable qu’en utilisant des critères strictement cliniques. (16)

Its short-term use may prove beneficial: (36)

● After a change in treatment, to assess the response to therapy.
● When symptoms are pronounced, to confirm the extent of respiratory impairment.
● To help identify aggravating factors at home (e.g., rooms with mold or dust). Its long-term use may prove beneficial:
● For early detection of exacerbations (in patients with few symptoms) and early activation of the crisis action plan.
● In patients with severe exacerbations.
● In patients with severe asthma (difficulty controlling asthma).

There may be a 20% variation in peak flow readings. In this case, three exhalations should be taken each time the device is used, and the highest value should be recorded.

Its use can aid in the diagnosis of asthma (if clinical findings are consistent with asthma and spirometry is not available); normal DEP values do not rule out a diagnosis of asthma.

Improvements in peak flow readings are generally observed within 3 months of starting inhaled corticosteroids. To ensure an adequate response to treatment, patients are asked to take two measurements per day for 2 weeks and record them in a chart. Significant variability in peak flow readings indicates uncontrolled asthma and a higher risk of an asthma attack. (35)

Peak expiratory flow rate standards are based on height in pediatrics. Peak expiratory flow rate can be estimated using apps. (37)

Tracking peak flow values in relation to treatment administration can be plotted on a graph, allowing for an assessment of asthma control and the need for treatment adjustments. (38)

One potential area for improvement could be the use of the peak expiratory flow in pediatric emergency departments to assess severity upon arrival, as well as to guide the decision to discharge patients treated in the emergency department or to help determine whether hospitalization is indicated. To achieve this, standardizing its use and providing training for medical and paramedical staff is crucial.

#### 4.3.3 Using a pulse oximeter at home

A pulse oximeter is a noninvasive, easy-to-use device that measures blood oxygen saturation. It is widely used in medical and paramedical settings, but its usefulness in the homes of patients with asthma has not yet been proven..

Although oxygen saturation—when it is 94% or lower—is one of the criteria used to assess the severity of an asthma attack, there are several limitations to the optimal use of a pulse oximeter:

➔ poor perfusion at the measurement site.

➔ irregular pulse.

➔ skin color and thickness.

➔ incorrect positioning.

➔ the use of adult pulse oximeters on children.

➔ devices have a margin of error of +/-4% for saturation levels between 70% and 100%.

➔ decreased oxygen saturation in children after taking SABA.

➔ interpreting saturation outside the context of a crisis.

All of these confounding factors, when considered in isolation, can lead to a misinterpretation of the severity of an asthma attack and, consequently, increase anxiety among caregivers and lead to more frequent visits to the emergency room.

It can be assumed that appropriate use of the device at home by patients must be accompanied by a good understanding of the instructions for use and a solid knowledge of asthma.

A study conducted in 2015 examined the use of pulse oximeters at home by patients with asthma, and the results were inconclusive. (39)

### 4.4 Communication and prevention

#### 4.4.1 Public health interventions

Asthma awareness and prevention must begin on a small scale with healthcare professionals but must also be part of the public discourse.

To this end, the Ministry of Health implemented an action program for asthma prevention and management from 2002 to 2005 with specific objectives.

The regional health agency must be made aware of the challenges surrounding asthma awareness and prevention, as well as the shortcomings of our current healthcare system. Thus, funds could be allocated to create prevention positions (such as strengthening asthma education teams in schools), as well as to support the development and reorganization of healthcare structures.

At the local level, outreach campaigns could be organized in schools (through school health programs, for example), on the radio, on TV, or in newspapers.

#### 4.4.2 Develop print and digital patient information materials

The sharing of information upon discharge from the pediatric emergency department varies depending on the healthcare providers involved and the specific work environment.

However, communicating with parents before the child returns home is important to help them understand the care plan that has been put in place. (34) To this end, a comprehensive informational resource could be developed and systematically provided to patients with asthma before they return home. (40) It could consist of:

❖ a printed action plan.

❖ measures to prevent exposure to and raise awareness of allergens and irritants (tobacco).

❖ criteria for assessing the severity of an asthma attack.

❖ a list of useful phone numbers (pulmonologists, asthma education program, EFR office, emergency medical services).

❖ video links to help with the use of inhalers.

❖ an informational brochure on the benefits of asthma school.

❖ a written notice for a close-monitoring follow-up appointment.

❖ digital tools available.

One area for improvement would be the use and evaluation of available digital tools. Currently, several apps exist to help manage asthma (and are available on smartphones). For example, the “findair” app helps track medication use and alerts users to poor asthma control if rescue medication is used too frequently. “Smart Asthma” allows users to monitor lung function using a peak flow meter. Some peak flow meters are equipped with Bluetooth or connect directly to a smartphone, allowing users to enter readings directly and facilitating monitoring. The “effic’asthme” app was developed by the AP-HP in 2019. It is a virtual asthma attack simulation app that allows parents and patients to practice responding appropriately to an asthma attack.

#### 4.4.3 Expand the asthma school

In our study, a small proportion of parents had heard of the asthma school, and only 15.3% had taken their child for a consultation there.

A study conducted at the CHUM from January through December 2018 (the EDELMA study) documented significant changes in habits as well as positive changes in medication adherence and healthcare utilization—particularly visits to the emergency department—among patients enrolled in the asthma education program. Awareness and prevention sessions were held weekly at the CHUM, and individual consultations were offered to promote treatment adherence. During these sessions, several topics were addressed, such as empowering the child, recognizing the signs of an asthma attack, communication between children and parents, inhalation techniques, and various other prevention strategies. This opportunity for discussion proved to be essential. Among the children attending the asthma clinic in this study, only 2.4% were referred by their primary care physician (66% were referred from the emergency department and 27% from the inpatient unit). This trend appears to be reversing at present, which may point to a lack of communication in the emergency department.

A medical thesis conducted in Martinique in 2015, which described the practices of general practitioners in the region, found that 83% of the physicians surveyed were familiar with the “Asthma School.” Rather than a lack of knowledge, a lack of familiarity with its use in daily practice may be suspected. A questionnaire was distributed to each physician (240 practitioners), and the response rate was 37%. The objective of the study was to evaluate the management of asthma by general practitioners in the territory and to compare it to the guidelines that had been in effect for 10 years at that time. In this thesis, 82% of the physicians surveyed wanted to be able to delegate therapeutic education to a facility or a trained professional, while 72% felt they were not providing enough therapeutic education. However, therapeutic education is key to asthma management and is central to the work carried out by the asthma school.

Currently, the asthma school limits its activities to pediatrics, but the expansion of this program (in terms of staff and resources) could facilitate the extension of its services to adults. A better understanding of the disease in adults could thus have a beneficial effect on the care of children.

#### 4.4.4 Expand home-based prevention interventions

Currently, a project to assess homes for irritants and allergens is being developed in collaboration with the Asthma School. This type of field intervention could facilitate research, concrete action, and awareness-raising among families. With the families’ consent, it could also enable health authorities in Martinique to collect and analyze data in order to promote behaviors aimed at combating and controlling allergens (preventive measures), as well as to take direct action in unsanitary homes (such as pest control, rodent control, and mold remediation).

## CONCLUSION

Our study shows that prehospital care for asthma attacks is inadequate for the majority of patients and that there is still room for improvement. Nevertheless, the figures on the use of therapeutic resources are quite similar to—and in some cases even better than—those in studies conducted in metropolitan France, suggesting that the efforts of medical staff are not in vain. Areas for improvement were identified throughout this study with the aim of enhancing the overall care of children with asthma in Martinique. One of our future objectives will be to develop a joint action plan and implement preventive measures within the MFME’s pediatric emergency departments. We also believe that our score could benefit from being studied in a prospective cohort to validate—or refute—its usefulness.

## Data Availability

All data produced in the present study are available upon reasonable request to the authors

## Supplementary data

**Supplementary table 1:** PRAM score for assessing the severity of an asthma attack.

| **Severity** | **PRAM Score** |  |  |  |
|---|---|---|---|---|
| Mild | 1-3 |  |  |  |
| Moderate | 4-7 |  |  |  |
| Severe/Life-Threatening | 8-12 |  |  |  |
| Signs | 0 | 1 | 2 | 3 |
| Suprasternal indrawing | Absent |  | Present |  |
| Scalene muscle contractions | Absent |  | Present |  |
| Wheezing | Absent | Expiratory only | Expiratory +/- inspiratory | Audible wheeze / silent chest / minimal air entry |
| Air entry | Normal | Decreased at bases | Widespread decreased |  |
| Room air O2 saturation | >94% | 92-94% | <92% |  |

**Supplementary 1:** Questionnaires distributed in the pediatric emergency department for children of age 6 years old or older

### Factors motivating a visit to the emergency room

● Have you ever felt that your child was in danger? Yes/No

● Did you feel helpless in the face of the situation (loss of control)? Yes/No

● Did you feel like you had tried every possible therapeutic approach at home? Yes/No

● Have you seen a doctor for this asthma attack within the past 5 days? Yes/No

### Medical follow-up of the child

● Is your child’s asthma being monitored by a doctor? Yes/No

○ If so, who is treating him/her (one or more answers possible):
○ Primary care physician / Pulmonologist / Pediatrician / Allergist / Other: …
● At what age did your child receive their first prescription for Ventolin: (Approximately)
● When was your last appointment with the doctor treating your asthma?

○ 0 to 6 months / 6 months to 1 year / over 1 year
● Has your child ever undergone a pulmonary function test? Yes/No

○ If so, what year was the most recent pulmonary function test conducted?
● Does your child have an individualized education plan at school?

○ If so, who did it?

### Asthma management

● Over the past 4 weeks, has your child had asthma-related symptoms lasting several minutes, more than 2 days a week? Yes/no

● Over the past 4 weeks, has your child’s usual activities been limited due to asthma? Yes/No

● Over the past 4 weeks, how many times has your child used a rescue inhaler or received nebulizer treatment (such as Ventolin)?

Less than two times a week / Two times or more a week

● Over the past 4 weeks, has your child woken up during the night because of his or her asthma? Yes/No

### History of asthma

● Does your child have any allergies—food, dust mites, dogs, cats, or grasses? Yes/No/Others

● Has your child received oral corticosteroid treatment in the past 6 months

● (Solupred / Celestene)? Yes/No/Don’t know

● Has your child ever been hospitalized in the intensive care unit for an

● asthma attack before? Yes/No

### Parents’ Knowledge

● What long-term asthma treatment is your child currently taking (treatment that he or she takes every day)?

Flixotide / Seretide / Singulair / I don’t know / Other …

● Have you heard of asthma school? Yes/No

● Have you attended an education class at asthma school (at least one parent)? Yes/No

● Do you think you know how to use an inhaler at home to take Ventolin? Yes/No

● Do you keep a crisis calendar? Yes/No

● Do you have a peak flow meter at home to assess the severity of your child’s asthma attacks? Yes/No

● Do you think you know how to use it? Yes/No

### Home-delivered medications for the current crisis

● Do you have a printed asthma action plan at home or within easy reach (a plan that specifies which medication to administer during an asthma attack, as well as the frequency and dosage)? Yes/No

○ If so, did you follow the action plan before coming to the emergency room? Yes/No

● Was this plan given to you in the pediatric emergency department? Yes/No

● Do you have a valid prescription for Ventoline (or Ventoline itself) at home? Yes/No

● What treatments were administered before arrival at the emergency room? Yes/No Ventolin / Corticosteroids (Solupred / Celestene) / None / Other …

● How many consecutive puffs of Ventolin did you give for each dose before you arrived at the emergency room (equivalent to 1 series)? *Circle your answer*.

0 - 1 - 2 - 3 - 4- 5 - 6 - 7 - 8 - 9 -10

● How long did you wait between each Ventolin puff (the time you waited before starting a new series) before you arrived at the emergency department?

<20 min / 20 min to 1 hr / 1 hr to 4 hrs / >4 hrs

● How many times did you take your Ventolin inhaler before arriving at the emergency department?

1 time / 2 times / 3 times / Other …

● If you use an inhaler chamber, how many breaths do you take, or how long do you hold it after each puff of Ventolin?

● How much time elapsed between the first dose of Ventolin and arrival at the emergency room? (Or, from the onset of symptoms to arrival at the emergency room if no Ventolin was prescribed)

<2 hours / between 2 and 6 hours / between 6 and 24 hours / >24 hours

● Did you call the emergency medical services for advice before coming to the emergency department? Yes/No

**Supplementary figure 1:**
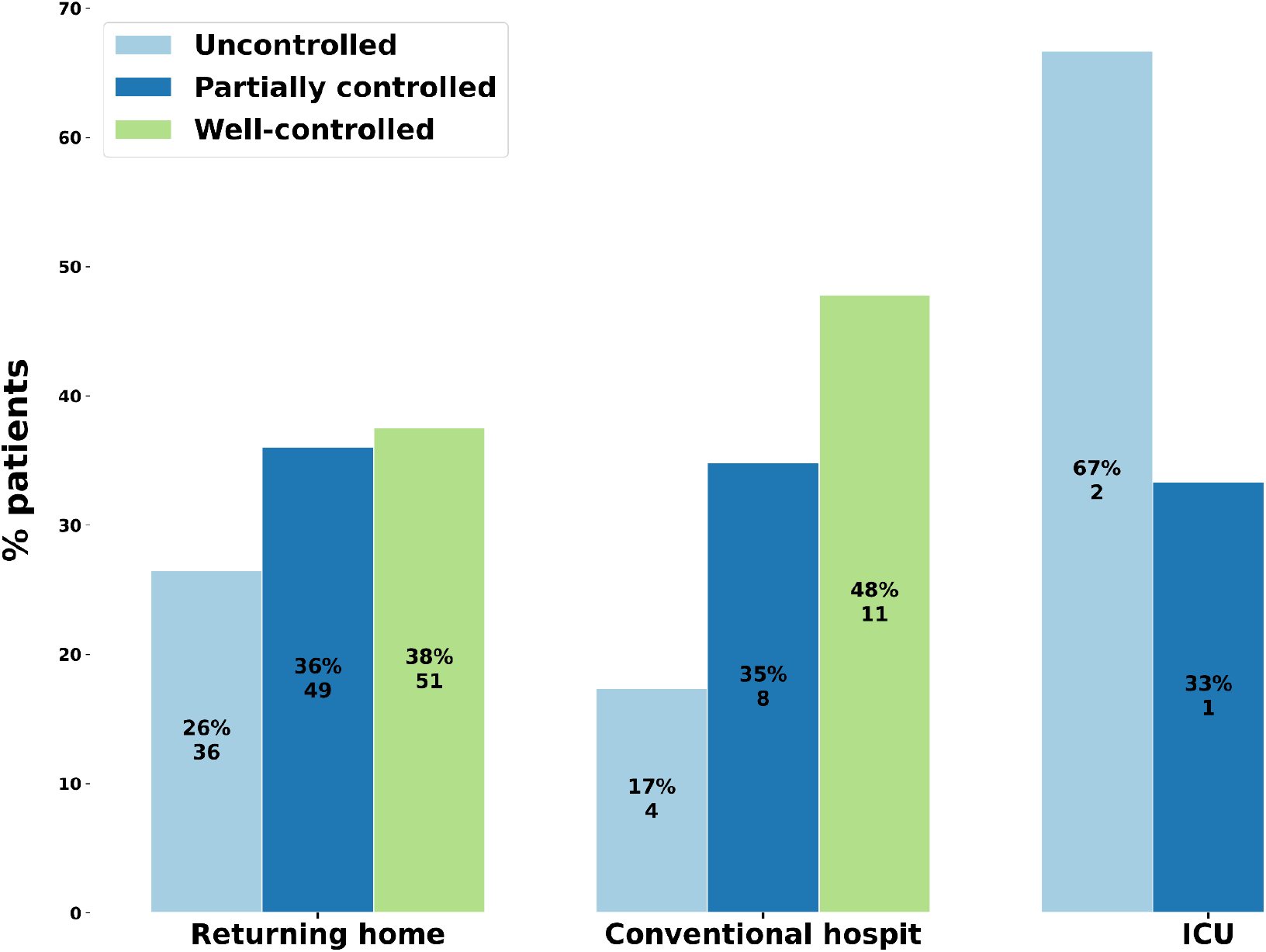
Patient discharge location based on asthma control in the emergency department. ICU: Intensive care unit, hospit: hospitalization

**Supplementary figure 2:**
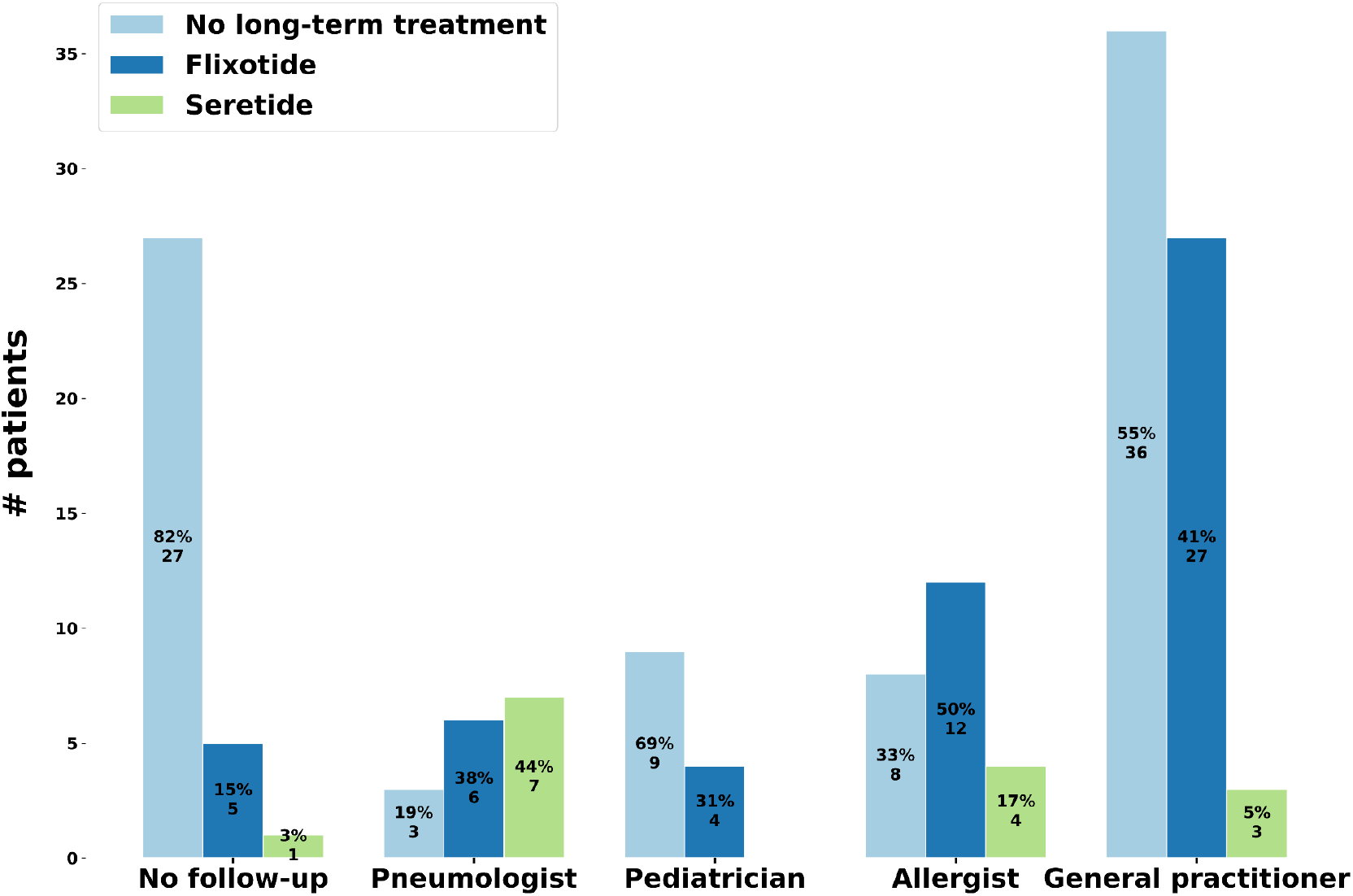
Type of long-term treatment based on medical follow-up by specialty.

**Supplementary figure 3:**
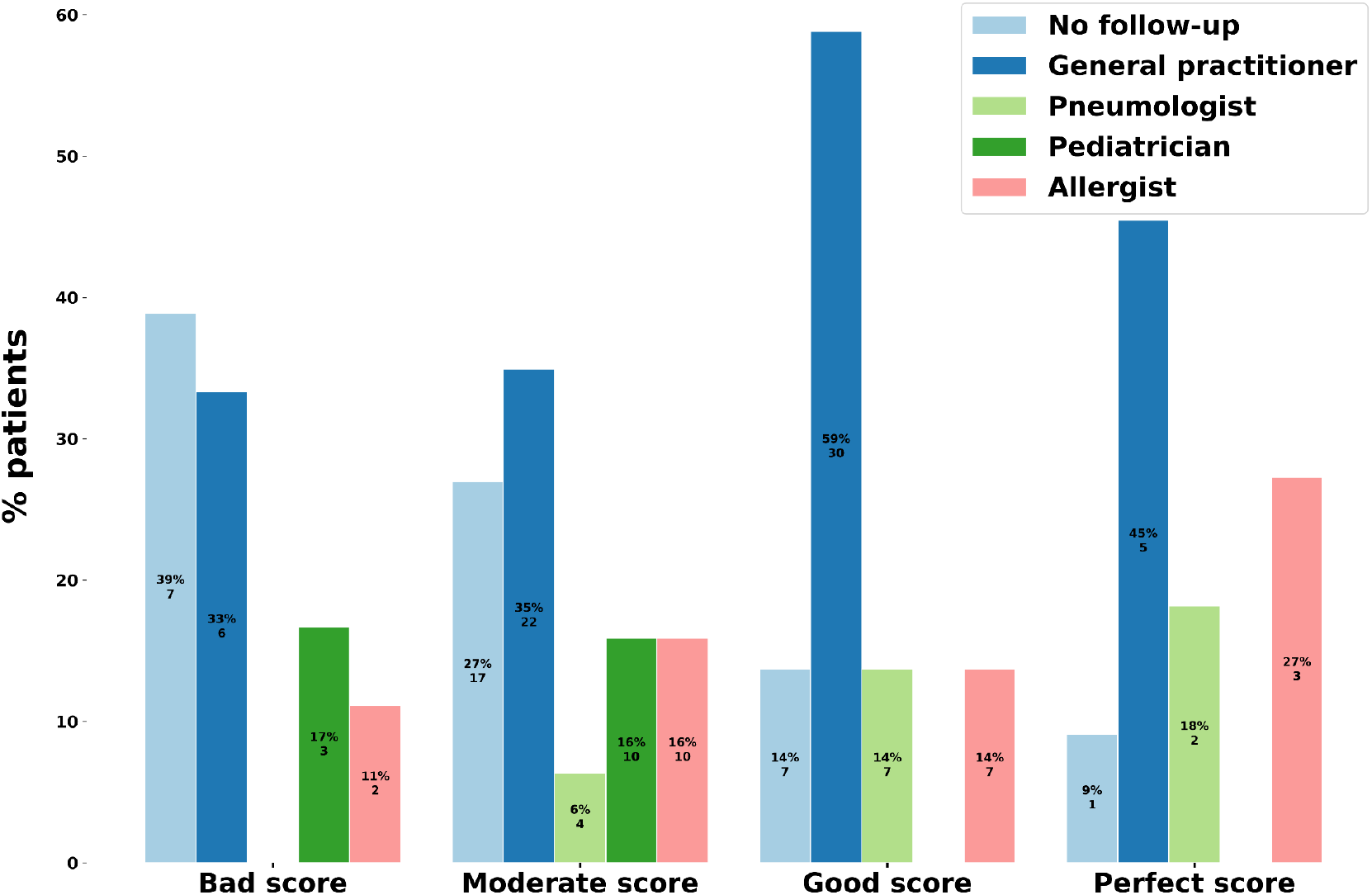
Therapeutic management of asthma attacks score Based on Medical Follow-Up.

**Supplementary figure 4:**
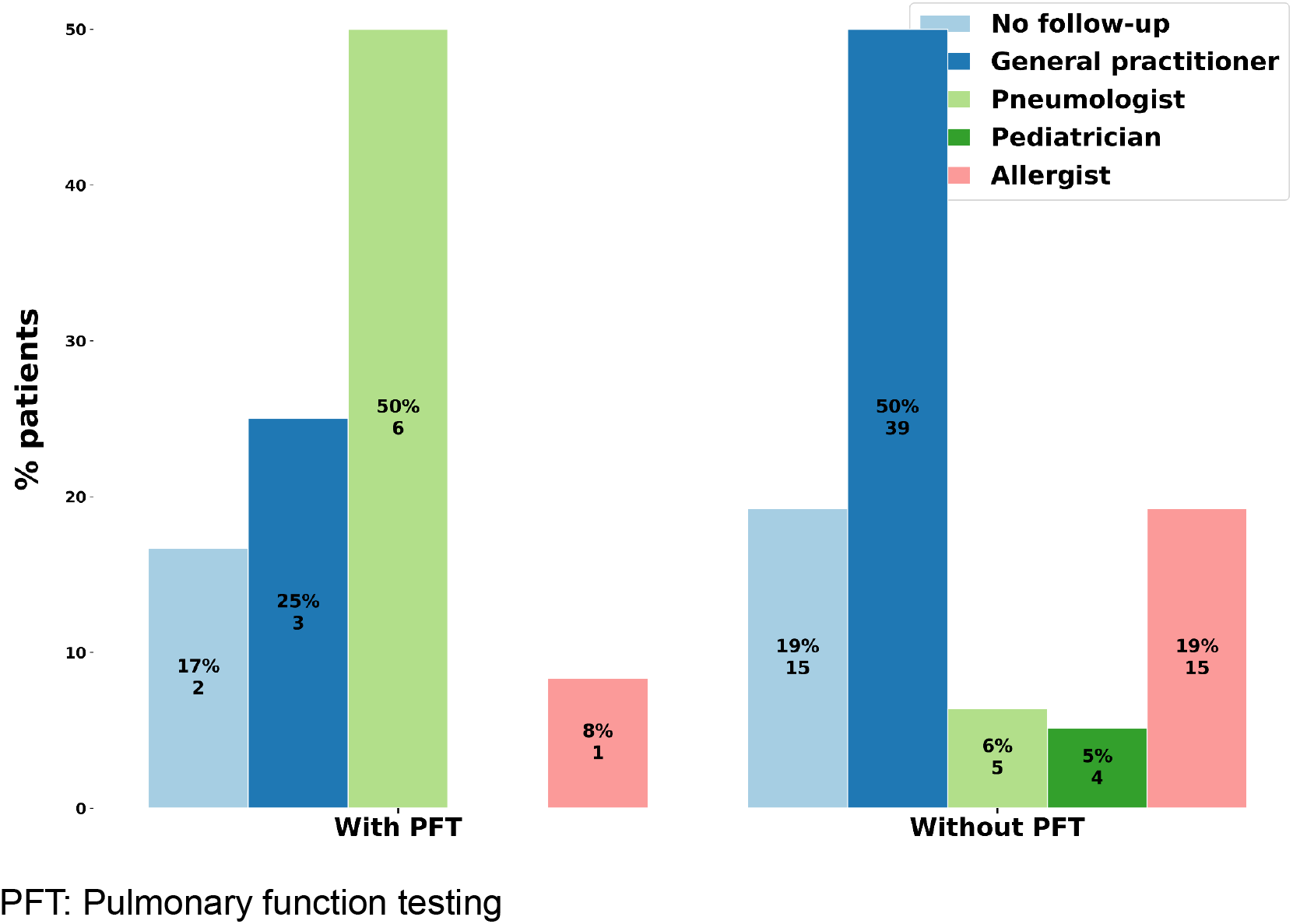
Conducting pulmonary function testing for children over 6 years of age as part of their medical follow-up.

**Supplementary figure 5:**
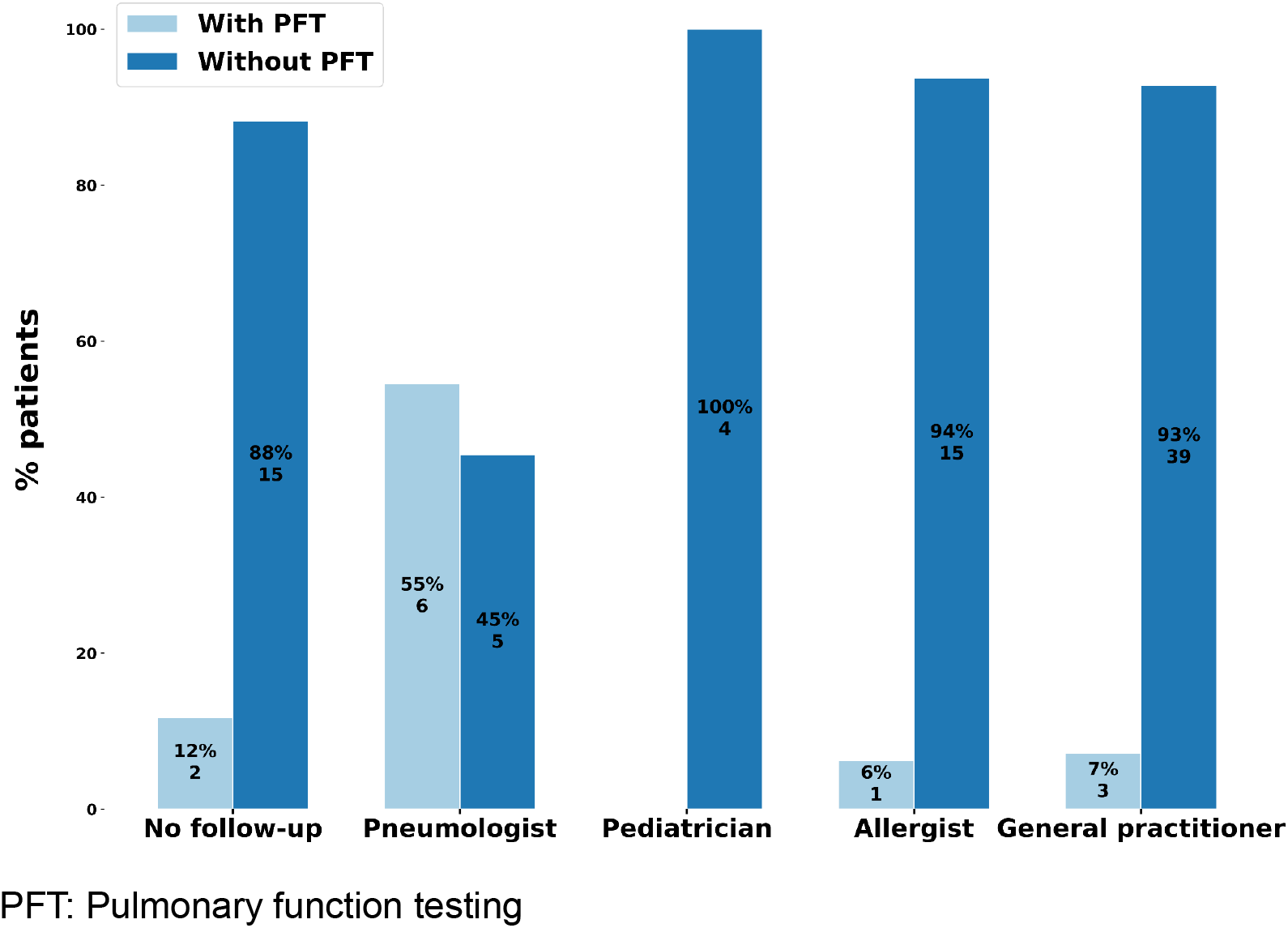
Conducting pulmonary function testing for children over 6 years of age as part of their medical follow-up.

## BIBLIOGRAPHIE

1. Asthme [Internet]. [cité 22 oct 2024]. Disponible sur: https://www.who.int/fr/news-room/fact-sheets/detail/asthma

2. Inserm [Internet]. [cité 22 oct 2024]. Asthme · Inserm, La science pour la santé. Disponible sur: https://www.inserm.fr/dossier/asthme/

3. Asthme [Internet]. [cité 21 août 2024]. Disponible sur: https://www.santepubliquefrance.fr/maladies-et-traumatismes/maladies-et-infections-respiratoires/asthme

4. Rivoyre RT de. Effets à court terme des poussières de sable du Sahara sur les consultations pour exacerbation d’asthme aux urgences pédiatriques du CHU de Pointe-à-Pitre (en 2011). 14 avr 2016 [cité 29 sept 2024];65. Disponible sur: https://dumas.ccsd.cnrs.fr/dumas-01364700

5. Wenzel SE. Asthma phenotypes: the evolution from clinical to molecular approaches. Nat Med [Internet]. mai 2012 [cité 1 nov 2024];18(5):716-25. Disponible sur: https://www.nature.com/articles/nm.2678

6. Ducharme FM, Zemek R, Chauhan BF, Gravel J, Chalut D, Poonai N, et al. Factors associated with failure of emergency department management in children with acute moderate or severe asthma: a prospective, multicentre, cohort study. The Lancet Respiratory Medicine [Internet]. 1 déc 2016 [cité 12 nov 2024];4(12):990-8. Disponible sur: https://www.sciencedirect.com/science/article/pii/S2213260016301606

7. Asthme [Internet]. SP2A. [cité 22 oct 2024]. Disponible sur: https://www.sp2a.fr/spa_documentation/asthme/

8. Westerhof GA, Coumou H, de Nijs SB, Weersink EJ, Bel EH. Clinical predictors of remission and persistence of adult-onset asthma. J Allergy Clin Immunol. janv 2018;141(1):104–109.e3.

9. Masson E. EM-Consulte. [cité 22 oct 2024]. Qualité de vie dans l’asthme chez l’enfant et ses parents, nouveau concept à intégrer dans l’évaluation du niveau de contrôle de la maladie. Disponible sur: https://www.em-consulte.com/article/1657605/qualite-de-vie-dans-l-asthme-chez-l-enfant-et-ses-

10. Tai A, Tran H, Roberts M, Clarke N, Wilson J, Robertson CF. The association between childhood asthma and adult chronic obstructive pulmonary disease. Thorax. sept 2014;69(9):805-10.

11. Webmaster S. Société de Pneumologie de Langue Française. 2024 [cité 1 nov 2024]. Recommandation de la SP2A prise en charge l’asthme de l’enfant de 6 à 12 ans. Disponible sur: https://splf.fr/recommandation-de-la-sp2a-prise-en-charge-lasthme-de-lenfant-de-6-a-12-a ns/

12. Suissa S, Ernst P, Kezouh A. Regular use of inhaled corticosteroids and the long term prevention of hospitalisation for asthma. Thorax. 2002;57(10):880-4.

13. Broquet Ducret C, Verga ME, Stoky-Hess A, Verga J, Gehri M. Impact d’une école de l’asthme sur la consommation en soins et la qualité de vie des enfants âgés de 4 à 12 ans et de leurs parents. Archives de Pédiatrie [Internet]. 1 nov 2013 [cité 14 nov 2024];20(11):1201-5. Disponible sur: https://www.sciencedirect.com/science/article/pii/S0929693X13004739

14. Observatoire des territoires - ANCT - Indicateurs : cartes, données et graphiques [Internet]. [cité 19 oct 2024]. Disponible sur: https://www.observatoire-des-territoires.gouv.fr/outils/cartographie-interactive/#c=indicator&f=medg&i=offre_sante_nombre.nombre&s=2023&selcodgeo=972&view=map9

15. Trousseau de poche [Internet]. [cité 14 oct 2024]. Disponible sur: https://trousseaudepoche.fr/

16. GINA-2024-Strategy-Report-24_05_22_WMS.pdf [Internet]. [cité 22 août 2024]. Disponible sur: https://ginasthma.org/wp-content/uploads/2024/05/GINA-2024-Strategy-Report-24_05_22_WMS.pdf

17. Asthme [Internet]. [cité 23 août 2024]. Disponible sur: https://www.santepubliquefrance.fr/maladies-et-traumatismes/maladies-et-infections-respi ratoires/asthme

18. Deschildre A, Abou-Taam R, Drummond D, Giovannini-Chami L, Labouret G, Lejeune S, et al. Mise à jour des recommandations (2021) pour la prise en charge et le suivi des patients asthmatiques adolescents (de 12 ans et plus) sous l’égide de la Société de pneumologie de langue française (SPLF) et de la Société pédiatrique de pneumologie et allergologie (SP2A). Version longue. Revue des Maladies Respiratoires [Internet]. 1 févr 2022 [cité 22 sept 2024];39(2):e1-31. Disponible sur: https://www.sciencedirect.com/science/article/pii/S0761842521003338

19. Inserm [Internet]. [cité 8 nov 2024]. Asthme · Inserm, La science pour la santé. Disponible sur: https://www.inserm.fr/dossier/asthme/

20. Pamuk G, Aboutaam R, Le Bourgeois M, De Blic J, Delacourt C, Lezmi G. Coût de santé de l’asthme sévère de l’enfant : évaluation individuelle du coût total. Revue Française d’Allergologie [Internet]. 1 avr 2019 [cité 14 nov 2024];59(3):293. Disponible sur: https://www.sciencedirect.com/science/article/pii/S1877032019301757

21. Nwaru BI, Ekström M, Hasvold P, Wiklund F, Telg G, Janson C. Overuse of short-acting β2-agonists in asthma is associated with increased risk of exacerbation and mortality: a nationwide cohort study of the global SABINA programme. Eur Respir J. avr 2020;55(4):1901872.

22. Stanford RH, Shah MB, D’Souza AO, Dhamane AD, Schatz M. Short-acting β-agonist use and its ability to predict future asthma-related outcomes. Ann Allergy Asthma Immunol. déc 2012;109(6):403-7.

23. de Blic J, Boucot I, Pribil C, Robert J, Huas D, Marguet C. Control of asthma in children: still unacceptable? A French cross-sectional study. Respiratory Medicine [Internet]. 1 sept 2009 [cité 29 oct 2024];103(9):1383-91. Disponible sur: https://www.sciencedirect.com/science/article/pii/S0954611109000870

24. Kaiser SV, Huynh T, Bacharier LB, Rosenthal JL, Bakel LA, Parkin PC, et al. Preventing Exacerbations in Preschoolers With Recurrent Wheeze: A Meta-analysis. Pediatrics [Internet]. 1 juin 2016 [cité 29 oct 2024];137(6):e20154496. Disponible sur: 10.1542/peds.2015-4496

25. Benkahoul Y, Bouchair N. Effet des corticoïdes inhales sur la croissance staturale des enfants asthmatiques ages de 6 à 12 ans suivis en consultation dans un service de pédiatrie générale [Internet] [Thesis]. Université Constantine 3 Salah Boubnider, Faculté de médecine; 2024 [cité 29 sept 2024]. Disponible sur: http://localhost:8080/xmlui/handle/123456789/5833

26. Julian V, Pereira B, Labbé A, Amat F. Caractéristiques des consultations pour exacerbation d’asthme aux urgences pédiatriques. Évaluation et perspectives pour une amélioration de la gestion pré-hospitalière. Revue des Maladies Respiratoires [Internet]. 1 janv 2014 [cité 29 oct 2024];31(1):13-20. Disponible sur: https://www.sciencedirect.com/science/article/pii/S0761842513001587

27. Blanc FX, Postel-Vinay N, Boucot I, De Blic J, Scheinmann P. The AIRE Study: Data analysis of 753 European children with asthma. Revue des Maladies Respiratoires. 2002;19(5 I):585-92.

28. Kergommeaux CC de. Vécus et représentations de la maladie asthmatique chez les parents d’enfants souffrant d’asthme à Mayotte. 22 août 2018 [cité 29 oct 2024];54. Disponible sur: https://dumas.ccsd.cnrs.fr/dumas-01903095

29. Schatz M, Zeiger RS, Yang SJ, Chen W, Crawford W, Sajjan S, et al. The relationship of asthma impairment determined by psychometric tools to future asthma exacerbations. Chest. janv 2012;141(1):66-72.

30. Hiscock H, Perera P, Mclean K, Roberts G, Lucas G, Kelly M, et al. Variation in paediatric clinical practice: A review of care in inpatient, outpatient and emergency department settings. Journal of Paediatrics and Child Health [Internet]. 2016 [cité 10 nov 2024];52(7):691-3. Disponible sur: https://onlinelibrary.wiley.com/doi/abs/10.1111/jpc.13212

31. Chalumeau M, Salanave B, Assathiany R, Kemeny J, Breart G. Connaissance et application par des pédiatres de ville de la conférence de consensus sur les rhinopharyngites aiguës de l’enfant. Archives de Pédiatrie [Internet]. 1 mai 2000 [cité 29 oct 2024];7(5):481-8. Disponible sur: https://www.sciencedirect.com/science/article/pii/S0929693X0089003X

32. Tavender E, Babl FE, Middleton S. Review article: A primer for clinical researchers in the emergency department: Part VIII. Implementation science: An introduction. Emergency Medicine Australasia [Internet]. 2019 [cité 29 oct 2024];31(3):332-8. Disponible sur: https://onlinelibrary.wiley.com/doi/abs/10.1111/1742-6723.13296

33. Kumar A, Husk K, Simpson R, Johnson GD, Burton C. Use of emergency departments by children and young people following telephone triage: a large database study. Emerg Med J [Internet]. 1 nov 2024 [cité 11 nov 2024];41(11):668-75. Disponible sur: https://emj.bmj.com/content/41/11/668

34. Curran JA, Gallant AJ, Zemek R, Newton AS, Jabbour M, Chorney J, et al. Discharge communication practices in pediatric emergency care: a systematic review and narrative synthesis. Syst Rev [Internet]. 3 avr 2019 [cité 11 nov 2024];8(1):83. Disponible sur: 10.1186/s13643-019-0995-7

35. Feldman JM, Rastogi D, Warman K, Serebrisky D, Arcoleo K. Peak Flow Feedback Intervention Improves Under-Perception of Airflow Limitation in Pediatric Asthma: A Randomized Clinical Trial. Annals ATS [Internet]. 25 oct 2024 [cité 29 oct 2024]; Disponible sur: https://www.atsjournals.org/doi/abs/10.1513/AnnalsATS.202406-637OC

36. Cowie RL, Revitt SG, Underwood MF, Field SK. The effect of a peak flow-based action plan in the prevention of exacerbations of asthma. Chest. déc 1997;112(6):1534-8.

37. Medicalcul - Débit Expiratoire de Pointe pédiatrique (Peak-Flow) ∼ Pneumologie [Internet]. [cité 31 oct 2024]. Disponible sur: http://medicalcul.free.fr/peakflowped.html

38. Reddel HK, Marks GB, Jenkins CR. When can personal best peak flow be determined for asthma action plans? Thorax. nov 2004;59(11):922-4.

39. Welsh EJ, Carr R. Pulse oximeters to self monitor oxygen saturation levels as part of a personalised asthma action plan for people with asthma - Welsh, EJ - 2015 | Cochrane Library. [cité 6 oct 2024]; Disponible sur: https://www.cochranelibrary.com/cdsr/doi/10.1002/14651858.CD011584.pub2/full

40. Hannah R, Chavasse RJPG, Paton JY, Walton E, Roland D, Foster S, et al. Emergency department discharge practices for children with acute wheeze and asthma: a survey of discharge practice and review of safety netting instructions in the UK and Ireland. Archives of Disease in Childhood [Internet]. 1 juill 2024 [cité 25 juin 2024];109(7):536-42. Disponible sur: https://adc.bmj.com/content/109/7/536

